# Non-linear mendelian randomization: detection of biases using negative controls with a focus on BMI, Vitamin D and LDL cholesterol

**DOI:** 10.1101/2023.08.21.23293658

**Authors:** Fergus W Hamilton, David A Hughes, Wes Spiller, Kate Tilling, George Davey Smith

**Author notes:** Corresponding author: Fergus Hamilton, MRC Integrative Epidemiology Unit, University of Bristol, Oakfield House, Oakfield Road, BS8 2PS.

## Abstract

Mendelian randomisation (MR) is an established technique in epidemiological investigation, using the principle of random allocation of genetic variants at conception to estimate the causal linear effect of an exposure on an outcome. Extensions to this technique include non-linear approaches that allow for differential effects of the exposure on the outcome depending on the level of the exposure. A widely used non-linear method is the residual approach, which estimates the causal effect within different strata of the non-genetically predicted exposure (i.e. the “residual” exposure). These “local” causal estimates are then used to make inferences about non-linear effects. Recent work has identified that this method can lead to estimates that are seriously biased, and a new method - the doubly-ranked method – has been introduced as a possibly more robust approach. In this paper, we perform negative control outcome analyses in the MR context. These are analyses with outcomes onto which the exposure should have no predicted causal effect. Using both methods we find clearly biased estimates in certain situations. We additionally examined a situation for which there are robust randomised controlled trial estimates of effects - that of low density lipoprotein cholesterol (LDL-C) reduction onto myocardial infarction, where randomised trials have provided strong evidence of the shape of the relationship. The doubly-ranked method did not identify the same shape as the trial data, and for LDL-C and other lipids they generated some highly implausible findings. Therefore, we suggest that until there is extensive simulation and empirical methodological work demonstrating that these methods generally produce meaningful findings use of them is suspended. If authors feel it is imperative that they report results from them there should be strong justification for this, and a number of sanity checks (such as analysis of negative and positive control outcomes) should be provided.

## Introduction

Mendelian randomization (MR) is now a widely used epidemiological technique contributing to improved causal inference in settings in which unmeasured confounding may bias estimates [1–3]. MR findings have been consistent with randomised trial data in many settings [4], and have assisted causal inference as a component of triangulation of evidence [5,6] Recently, there have been developments in MR in which attempts are made to extend causal inference from a whole population (e.g. the overall effect of increasing vitamin D on all-cause mortality), to strata of that population (e.g. only those with low levels of vitamin D) [7–11]. This has the potential to answer important questions around the linearity of exposure-outcome effects. The development of the “residual” method led to MR studies that purported to show striking non-linear effects [12–18], in particular with respect to vitamin D and BMI as exposures. However, recent analyses have suggested that this method may be susceptible to serious bias, including that likely related to amplification of selection bias within strata [3,19–21]. In this manuscript, we explore a number of issues that suggest caution in using non-linear approaches at present.

### Development of the residual method and concerns about inconsistent causal estimates

In the residual method the measured levels of the exposure (e.g. Vitamin D) are regressed onto the genetic instrument (e.g. a polygenic risk score for Vitamin D). The residuals of that regression represent the IV-free exposure (sometimes referred to as the “non-genetic” portion of Vitamin D). These residuals are then split into strata (often 10, or 100 strata) ordered by the mean IV-free exposure value. Therefore, the sample in stratum 1 have the lowest level of “IV-free” Vitamin D, and the sample in stratum 10 (or 100) have the highest value. MR is then run within each stratum, and estimates are combined by parametric statistical methods (such as fractional polynomials) to estimate a non-linear curve. The requirement for stratification based on the residuals rather than the exposure itself is driven by the desire to avoid collider bias [22] as this would be induced by stratification on the exposure itself.

There has been a recent focus on the performance of this method. In particular, one study identified a non-linear predicted causal effect of Vitamin D on a number of outcomes, suggesting that increasing Vitamin D in those who were below the median level might produce considerable benefit (as opposed to the null overall causal effect) [23]. In some analyses, despite a precisely estimated null or harmful overall effect, every strata had a protective effect, with some effects substantial. Following the journal and the authors being alerted to this [19] the authors referred to it “as a logical impossibility if all estimates have a causal interpretation”[23] after the journal retracted the paper[24]. Several other analyses applying identical methods to the same exposure, with overlapping data and outcomes, have, unsurprisingly produced similar and likely equally spurious findings [15,16].

Following the documentation that the findings were erroneous the authors of the initial non-linear Vitamin D analysis [23] re-evaluated the residual method used in the study. In a recent paper [25] they showed via simulation studies that the residual method was biased under certain situations, in part relating to the “constant genetic effect” assumption – that the genotype-exposure effect is linear and thus constant across strata (i.e. the coefficient for IV-exposure remains the same across strata). For Vitamin D, the constant genetic effect assumption was shown not to be satisfied, with much larger genotype-exposure effect estimates in top strata than in the bottom strata [25]. They also noted that this heterogeneity in IV-exposure association was not identified in the residual method, and suggest that this is a flaw in the residual approach. It is not clear why this would have led to the J-shaped curve observed in the residual method, however. To address this challenge a different way of stratifying the exposure was developed, called the “doubly-ranked” method [11].

In this method, the strata are developed in multiple steps (**Figure 1**). In the first step, the population is doubly-ranked by the level of the genetic instrument into pre-strata. Subsequently, within each pre-stratum, the participant with the lowest level of the exposure is taken and placed into the lowest final stratum. The participant with the next lowest level of the exposure in pre-strata 1 is placed into final stratum two. The same process occurs for all pre-strata. The first stratum therefore contains the first participant of every pre-strata, each of which has the lowest level of exposure in the pre-stratum. The number of pre-strata (K) to be generated depends on the number of strata desired (J) and the total sample size (N) using the formula N = J x K. That is, for a sample size of 1,000, and to generate a final desired number of strata of 10, 100 pre-strata are generated.

**Figure 1:**
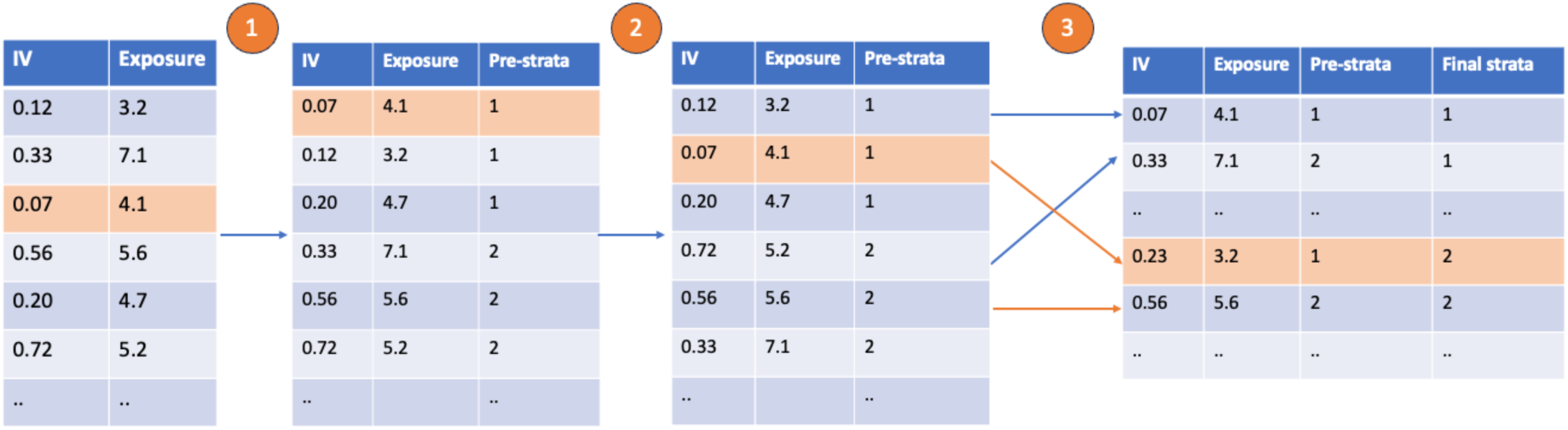
A worked example of strata generation in the doubly-ranked approach. In the first step, all participants are sorted by their level of the instrumental variable (IV) into pre-strata. In the second step, participants are sorted by their level of exposure within each pre-strata. In the third step, the first participant from each pre-strata is placed in the first final strata. The second participant in each pre-strata is placed in the second final strata, and so on. In the orange, a single participant is followed. This participant has the lowest level of IV so is placed in pre-strata 1. They have the second lowest level of exposure within this pre-stratum so are placed in the second final stratum.

A more detailed exposition is given elsewhere [11]. The doubly-ranked method is suggested (using simulation studies) by the authors to be insensitive to changes in the exposure to genotype association over strata, i.e. does not rely on the constant genetic effect assumption to generate unbiased estimates. The retracted non-linear vitamin D paper[18] – which claimed substantial non-linearity in the causal effect of vitamin D – has been replaced by a revised paper applying the doubly-ranked approach which suggests no meaningful non-linearity [23,24].

Despite the development of the doubly-ranked method solving some of the issues evident in both empirical and simulation studies of the residual method, there remain a number of uncertainties about the reliability of both methods to provide unbiased causal estimates. In this paper we evaluate the residual and doubly-ranked methods using “negative control outcome (NCO) analyses [5,6,26] in UK Biobank, a large and widely researched cohort study. NCO analyses are analyses performed on control outcomes that should lead to a null result, and where identification of a non-null estimate suggests bias. We additionally examined a situation for which there are robust randomised controlled trial estimates of effects - that of the effect of low density lipoprotein cholesterol (LDL-C) reduction on myocardial infarction, where randomised trials have provided strong evidence of the shape of the relationship [27,28]. We compared these estimates from trials with those generated by both non-linear MR methods. We present our findings in the context of a recent preprint from the principle developer of the doubly-ranked method, and draw attention to highly implausible findings in this report [29].

## Materials and Methods

### Data source

This study was performed in UK Biobank [30]. UK Biobank recruited around 500,000 participants between 2006-2010 from 22 sites across the UK. Participants were invited via post and had a range of interviews on recruitment to record previous life events, demographics, and medical history. Additionally, participants had blood samples taken for biochemical testing and had a range of physical and anthropometric measures performed (e.g. body mass index, blood pressure). Subsequently, participants had record linkage to electronic health record data for secondary care and national death data (for >99% of participants).

We analysed data on the subset of participants who were minimally related and of White British ancestry (n = 385,290), defined by relationship analyses [31]. Specifically, we used the MRC Integrative Epidemiology Unit genetically quality controlled data, described in detail elsewhere, excluding highly related participants [31].

### Exposures and data source

This study focussed on Vitamin D (measured as nmol/L) BMI (measured as kg/m^2^), and LDL-C and triglycerides (TG) (measured as mmol/L) as exposures, as all of these were measured in participants on recruitment to UK Biobank, and have been widely used in non-linear MR analyses [17,23,29,32]. Details on measurement are available via the UK Biobank website. Our study sample for each analysis included only participants who had these values recorded successfully on recruitment to UK Biobank and had genetic data available. Sample sizes differed slightly due to missing values for each exposure (e.g. due to assay failure, inability to give blood, etc). For triglycerides, we also extracted data from a subsequent visit, around 2 years later, for around 15,000 participants.

We then identified genetic variants to use in MR for all exposures. To identify variants associated with BMI, we used a large recent meta-analysis of BMI which did not include any participants of UK Biobank [33]. We extracted all variants which were associated with BMI (p < 5 x 10^-8^) and then clumped them by linkage disequilibrium to include only independent variants (r^2^ <0.001, kb = 10,000). LD clumping was performed using the *TwoSampleMR* package in R and was based on LD from the European ancestry participants from the 1000 Genome’s project [34]. In total we included 68 variants (**Supplementary Table S1**). For Vitamin D, to match as close as possible to a previously described non-linear MR analysis, we used the same set of variants that they used [23]. These have previously identified to be associated with Vitamin D levels in meta-analysis from four well characterised genomic regions known to be involved in Vitamin D metabolism (*GC, DHCR7, CYP2R1, CYP24A1*)[35]. In that analysis, 21 variants were used but 3 were unavailable in our data so in total 18 variants were used to generate our polygenic risk score (**Supplementary Table S2**). The three missing variants were all rare (minor allele frequency <0.005) in 1000 Genomes European subset) and were therefore unlikely to substantially affect our instrument. For LDL-C and TG we used summary statistics from the most recent analysis of the Global Lipids Genetics Consortium (GLGC) excluding UK Biobank participants. Again, we took independent variants (p < 5 x 10^-8^, r^2^ <0.001, kb = 10,000) from across the genome. In total we included 296 variants for LDL-C and 366 variants for TG (**Supplementary Table S3)**.

To generate an individual PRS for each participant we used PLINK v2.0.4 [36], with the summary effect generated by summed weighted allele scores, with each allele weighted by its effect on the exposure.

### Genotyping and imputation

This analysis was performed on quality controlled genetic data held within the MRC Integrative Epidemiology Unit. Full details of quality control are available elsewhere [31] but a summary is provided here. The full data release for this study contains the cohort of successfully genotyped samples (n=488,377). 49,979 individuals were genotyped using the UK BiLEVE array and 438,398 using the UK Biobank axiom array. Pre-imputation QC, phasing and imputation are described elsewhere [30]. In brief, prior to phasing, multiallelic SNPs or those with MAF ≤1% were removed. Phasing of genotype data was performed using a modified version of the SHAPEIT2 algorithm [37]. Genotype imputation to a reference set combining the UK10K haplotype and HRC reference panels was performed using IMPUTE2 algorithms [38]. The analyses presented here were restricted to autosomal variants using a graded filtering with varying imputation quality for different allele frequency ranges. Therefore, rarer genetic variants are required to have a higher imputation INFO score (INFO>0.3 for MAF >3%; INFO>0.6 for MAF 1-3%; INFO>0.8 for MAF 0.5-1%; INFO>0.9 for MAF 0.1-0.5%) with MAF and Info scores having been recalculated on an in-house derived ‘European’ subset. Data quality control Individuals with sex-mismatch (derived by comparing genetic sex and reported sex) or individuals with sex chromosome aneuploidy were excluded from the analysis (n=814).

### Degree of relatedness

Estimated kinship coefficients using the KING toolset identified 107,162 pairs of related individuals. An inhouse algorithm was then applied to this list and preferentially removed the individuals related to the greatest number of other individuals until no related pairs remain. These individuals were excluded (n=79,448). Additionally, 2 individuals were removed due to them relating to a very large number (>200) of individuals.

### Outcomes and covariates

Our primary outcomes for this study were age and sex for BMI and Vitamin D exposures, and myocardial infarction when LDL-C was the exposure. We chose age and sex as key negative control outcomes as these cannot be caused by the exposures in question and any non-null estimates in any strata suggest the analyses are biased [39]. However, there are well established selection effects relating to BMI, age, and sex into studies like UK Biobank (see e.g. [40]). It is plausible that these selection effects are amplified by non-linear methods and could lead to bias. This still, of course, means the apparent findings are unreliable.

Myocardial infarction was extracted from the UK Biobank algorithmic definition (Data Showcase field 42000). MI outcomes were defined as prevalent (occurring before study enrolment), incident (occurring after study enrolment), or both (including both incident and prevalent cases).

We extracted covariates including UK Biobank recruitment centre, smoking status (coded as current or never/ex) and the first 5 genetic principal components from UK Biobank directly. Smoking status was included for some sensitivity analyses given the known bidirectional association between smoking status and BMI [41].

### Negative control outcome analysis – BMI and Vitamin D

To perform our negative control outcome analysis we first performed conventional MR then used the *SUMnlmr* package in R to perform both the residual and doubly-ranked method on our individual level data [42]. Conventional MR was performed using a two stage residual inclusion approach in the *OneSampleMR* R package [43], using both the whole cohort and then in strata of age and sex. We first tested the effect of vitamin D and BMI (individually) on age and sex, firstly without covariates (unadjusted), and then age (when sex was an outcome), sex (when age was an outcome), and also including UK Biobank recruitment centre, and the first genetic five principal components. Non-linear MR was performed using the standard settings in the *SUMnlmr* package, with a Gaussian distribution for linear outcomes, and a binomial distribution for binary outcomes. We chose to use ten strata for most analyses; but performed sensitivity analyses using different numbers of strata.

For BMI, we performed an analysis stratified by smoking status as this had been performed as a key analysis in an as yet uncorrected high-profile paper using non-linear MR [17] As BMI has a bidirectional association with smoking, this could induce collider bias, but we repeated these analyses to determine if estimates were more biased when stratifying by smoking status.

### LDL cholesterol on myocardial infarction

In a further analysis, we focussed on the role of LDL-C as an exposure and myocardial infarction as an outcome. We provide the background to this positive control analysis in **Box 1**.

We performed two analyses. First, for completeness we replicated the negative control analysis discussed above (with age and sex as outcomes) using LDL-C as an exposure. Subsequently, we performed NLMR analyses on the effect of LDL-C on a) incident, b) prevalent, and c) all myocardial infarction to identify if effects matched randomised trial data.

As non-linear MR results could be altered by statins, which have a large effect on LDL-C levels, we performed sensitivity analyses in a cohort including only statin users, only non-statin users, and those under the age of 50 (where statin use was rare (∼5%) and therefore not likely to bias estimates). Finally, we also fitted a model adjusting for statins, age, sex, and the first five genetic principal components. We recognise that the approach of analysing only those who use/do not use statins or adjusting for statins generates collider bias[22] (as statin use is almost entirely downstream of LDL-C levels) but we included this for completeness.

#### Box 1

##### Purpose of positive control analysis and background to the relationship between LDL-C and MI

The purpose of positive control analyses is to perform an analysis whereby the true result is already known, and a comparison can be made to the results of a presented analysis and this truth. In this paper, we chose the effect of LDL-C on myocardial infarction.

We chose this example because this is a well-studied exposure outcome relationship, where conventional MR estimates match in direction to randomised trial estimates, although MR estimates are around 40% larger, presumably due to the lifelong effect of lipid reduction conferred by genotype compared to that of drugs [1,44,45]. In addition there are a large number of randomised trials with differing levels of baseline LDL-C [27]. These individual trial estimates have been meta-regressed to identify if the effectiveness of LDL-C reduction depends on the baseline LDL-C [27,28], so we can calculate the shape of the LDL-C and MI relationship.

In 2010, a meta-analysis of 26 trials (largely statin vs control) identified no clear evidence of the effect of baseline LDL-C on the effectiveness of LDL-C therapy (on all major vascular events), with a summary OR of 0.78 (95% CI 0.76-0.80) per 1mmol/L reduction in LDL-C [28]. In a more recent 2018 meta-analysis of 34 trials which reported specific LDL-C effects on myocardial infarction [27], the overall OR per 1 mmol/L LDL-C reduction was similar (0.76 95% CI 0.72-0.80), but in meta-regression they identified a small decrease in rate ratio (RR) of 0.90 (95% CI 0.84-0.97) with each 1 mmol/L increase in LDL-C (i.e. the effectiveness of therapy was greater in those with higher LDL-C levels). In those with an LDL-C of more than 4mmol/L, the OR per 1mmol/L reduction was 0.64 (95% 0.53-0.78), while in those trials with a baseline LDL-C of less than 2.5mmol/L, the OR was 0.85 (95% CI 0.76-0.92). The summed data therefore suggests a mostly linear effect with a moderate increase in effectiveness in those with *higher* LDL-C.

Given the number of trials across different settings and the overall sample size, we can be reasonably confident of the true shape of this relationship and can compare this estimate with that generated by other methods (e.g. non-linear MR).

### Triglycerides and cancer mortality

Finally, given a recent pre-print identifying biologically highly implausible results, we aimed to replicate an analysis of the effect of triglycerides on cancer mortality reported in the above preprint[29]. We used the same definitions of cancer mortality and performed non-linear MR adjusting for age, sex, age * sex, age*age, age*age*sex, and the first ten principal components. This choice of covariates was to match the prior analysis. We also – using the repeat sampling data available for around 15,000 participants – assessed the stability of strata for triglycerides, as these are known to be highly fluctuant. To do this we simply performed the doubly-ranked method at each time point (initial visit, repeat visit), and showed whether participants remained in the same strata using an alluvial plot.

In line with the authors of the doubly-ranked method recommendations [11] we performed multiple replicates (20) of each doubly-ranked approach and combined estimates using Rubin’s rule.

### Ethics

This study was performed under the UK Biobank application number 81499. UK Biobank was ethically approved by the North West Multi-centre Research Ethics Committee (MREC).

### Data and code availability

This analysis was performed in the UK Biobank. Data can be accessed via application to the UK Biobank. We provide code used to perform our analysis in a supplement.

## Results

### Negative control outcomes as an assessment of the potential risk of bias

For our negative control outcome analysis, we generated instrumental variables (IVs) for two exposures (Vitamin D and BMI) using a summed linear score across single nucleotide polymorphisms (SNPs) and performed conventional and non-linear MR on participant age in the self-reported white British population in UK Biobank.

In total, we included 351,005 participants in our analysis using Vitamin D as an exposure and 383,793 participants in our analysis on BMI. Demographics of the population are in **Supplementary Table S4**.

### Conventional MR estimates are close to the null

Conventional MR estimates of the effect of Vitamin D on age (beta per nmol/L: 0.0028; 95% CI - 0.004; 0.009, p = 0.39) and sex (OR per nmol/L 1.002; 95% CI 1.000 - 1.003, p = 0.06) were close to the null. MRs estimate of the effect of BMI on age (beta per kg/m2: -0.019, 95% CI -0.060 - 0.023, p = 0.37) and sex (OR per kg/m2 1.01; 95% CI 1.00 - 1.02, p = 0.04) were also close to the null.

We also calculated the effect of BMI and Vitamin D on age in males and females separately, and the effect of BMI and Vitamin D on sex in deciles of age. These are reported in **Supplementary Table S5**. Estimates across subgroups were similar – accepting the play of chance – across these analyses.

### Non-linear MR estimates diTer substantially across strata

We then used both the residual and doubly-ranked method to generate stratum-specific estimates of the effect of each exposure on age and sex in univariable analyses. We chose to use ten strata, although results were similar using a differing number of strata (data not shown).

In contrast to the null estimate across the whole cohort, estimates across each stratum were markedly different using either method (**Figure 2**). Focussing on the residual method, for the predicted effect of Vitamin D on age (**Figure 2A**), there were positive effect estimates in lower strata which decreased moving from lower to upper strata for the effect of Vitamin D on age, leading to null effects. However, the uppermost strata had a positive effect estimate. For BMI on age (**Figure 2C**), we saw a similar trend, with positive effect estimates in lower strata and negative effect estimates in higher strata.

**Figure 2:**
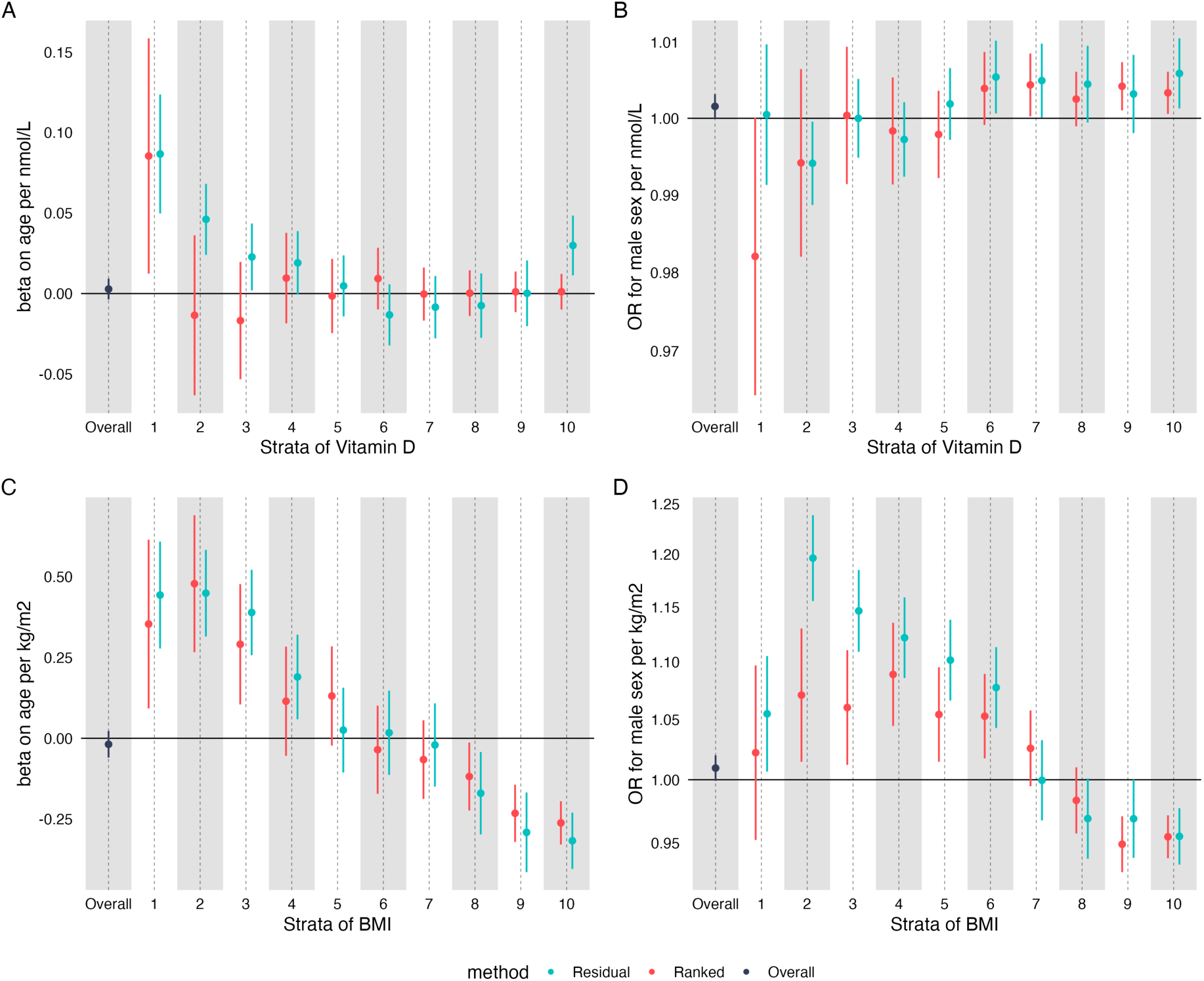
The predicted causal effect of Vitamin D on age (A), sex (B) and BMI on age (C) and sex (D). Estimates were generated in each stratum using the residual method (blue) and the doubly-ranked method (red) and were unadjusted for covariates. The black estimate represents the conventional MR estimate generated using the whole cohort.

For the effect of BMI on sex (**Figure 2D**), we saw positive estimates in lower strata and negative effects in upper strata, in line with a previous similar analysis we have performed, although that analyses used different parameters including a different instrumental variable[21]. In short the estimates would suggest that BMI increased the odds for being male in those with the lowest non-genetic BMI and increased the odds of being female in upper strata. For the effect of Vitamin D on sex (**Figure 2B**) we observed null effects in lower strata but modest positive effects in upper strata suggesting that vitamin D increases the odds of being male in those with the highest non-genetic BMI.

For the doubly-ranked method, we identified largely null estimates for the effect of Vitamin D on age (**Figure 2A**). For the effect of Vitamin D on sex (**Figure 2B**), effect estimates were similar between the residual and doubly-ranked method, with increasingly positive effect estimates when moving from lower to upper strata. For the effect of BMI on age (**Figure 2C**), we saw similar estimates to the residual method, with positive estimates in lower strata and negative estimates in upper strata. For the effect of BMI on sex (**Figure 2D**), we saw marked non-null estimates with a similar trend but, in general, estimates closer to the null for the doubly-ranked relative to the residual method. We calculated Cochran’s Q to formally assess the heterogeneity of strata specific estimates. These results are reported in **Supplementary Table S6**. There was evidence – extremely strong in many cases - of strata specific estimate heterogeneity across all negative control outcome analyses except the analysis of Vitamin D on age and sex using the doubly-ranked method.

Estimates were similar in analyses adjusted for covariates: age (for sex as an outcome), sex (for age as an outcome) and the first 5 genetic principal components (**Supplementary Figure S1**). Log-transforming Vitamin D (as recommended by the creators of the residual method) brought estimates from the residual model closer on average to the doubly-ranked model (**Supplementary Figure S2**), although these were still non-null across many strata.

For our BMI analysis, we then stratified by smoking status (as was performed in a previous non-linear MR analysis in BMI, producing the headline findings from the paper[17]) and re-ran analyses (**Supplementary Figure S3)**. Smoking had been shown to have a bidirectional relationship with BMI[41] before the non-linear *BMJ* MR paper was published[17], and therefore it should have been clear that this could have produced collider bias [22]. In these analyses, stratification on smoking made considerable difference to some strata-specific estimates, although the general shape of the association remained similar.

To summarise, using both the residual and doubly-ranked method we identified non-null, stratum-specific associations between two exposures (Vitamin D and BMI) and two negative control outcomes (sex and age) across strata of the exposure in which the expected result is null.

### LDL cholesterol and myocardial infarction

To examine both methods in a scenario where we anticipate the shape of the causal relationship we examined the association between LDL-C and a key outcome: myocardial infarction. RCT data from >30 trials have demonstrated strong and broadly linear effects of LDL-C reduction on both outcomes with meta-analyses identifying slightly larger effect estimates in those with higher levels of LDL-C at baseline, while a recent MR analysis identified the opposite effect (increased effectiveness of LDL-C reduction in those with lower LDL-C[29], see **Box 1** for further background).

First, as with analyses we presented above, we performed negative control outcome analyses of the effect of increasing LDL-C on age and sex. In conventional MR, estimates were essentially null for age (beta on age per 1mmol/L increase in LDL -0.10; 95% CI -0.23, 0.03, p = 0.15), but suggested increased LDL-C was ‘causal’ for being more female (OR 0.95; 95% CI 0.92-0.98, p = 0.004). Conventional MR estimates of the effect of LDL-C on age were similar in men and women (**Supplementary Table S6**), although there was some evidence of heterogeneity of MR estimates of LDL-C on sex across strata, p value for heterogeneity 0.03).

Compared to conventional MR estimates, divergence from the null was much greater in non-linear MR (**Figure 3**), with estimates on age as high as 5.74 (95% CI 1.79 - 9.63) years per 1mmol/L LDL-C increase in one strata from the doubly-ranked method, a ∼50 fold increase in bias compared to the conventional MR estimate. In contrast to our NCO analyses above, the bias was more extreme for the doubly-ranked method than the residual method.

**Figure 3.**
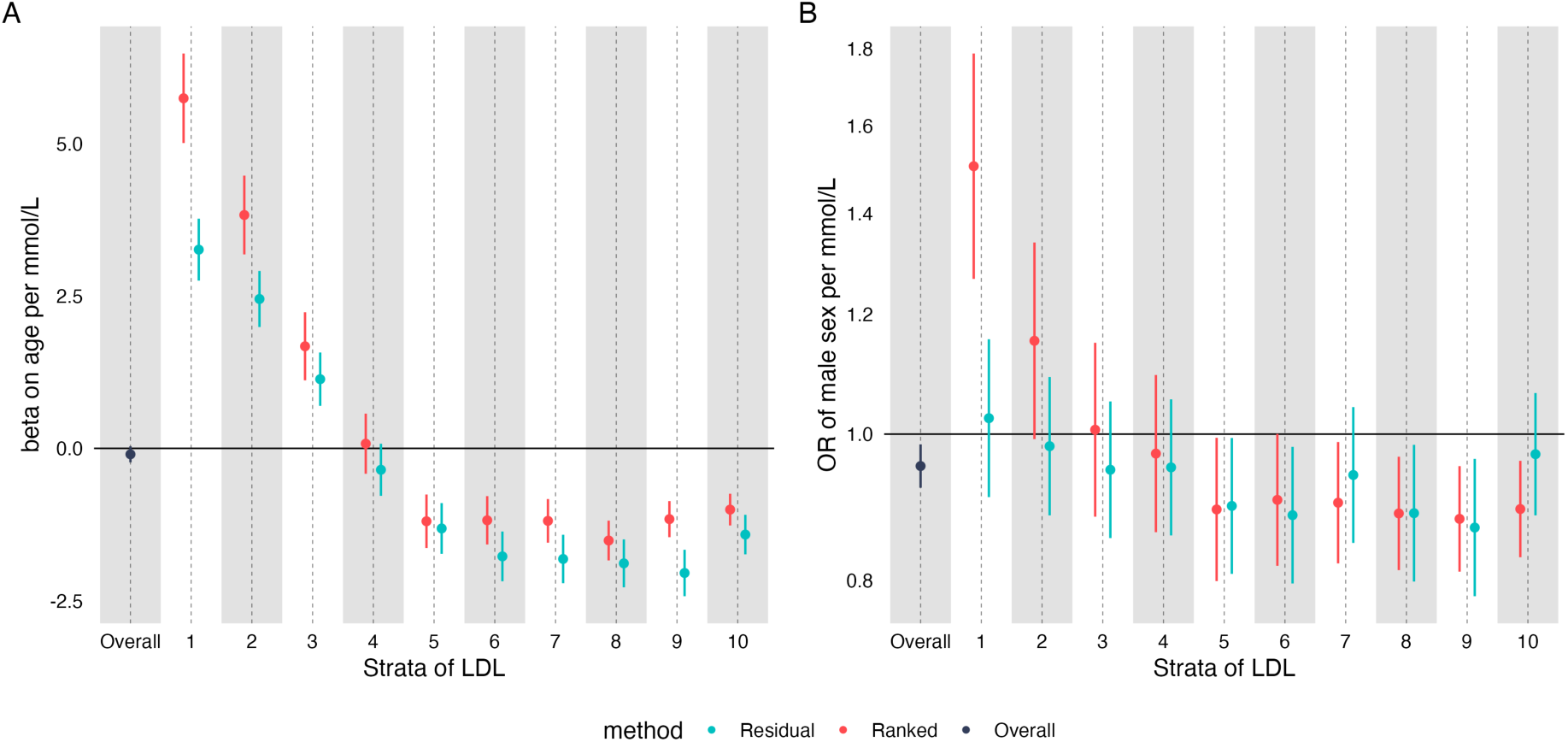
The predicted causal effect of increased LDL-C on (A) age and (B) sex. Estimates were generated in each stratum using the residual method (blue) and the doubly-ranked method (red) and were unadjusted for covariates.

We then went on to perform MR of the effect of increased LDL-C on myocardial infarction. For our primary analysis we included both incident and prevalent cases of MI, the effect on incident and prevalent MI separately are shown in **Supplementary Figure 4**. In conventional MR we saw the expected effect of LDL-C (OR for MI 1.74; 95% CI 1.61-1.87). However, in NLMR we saw unexpected effects, particularly with the doubly-ranked method. For the effect on MI (**Figure 4**), we saw large differences in effects across strata, with the strongest effect in those with the lowest LDL-C, and the weakest effects in those in strata 5, 6, and 8.

**Figure 4.**
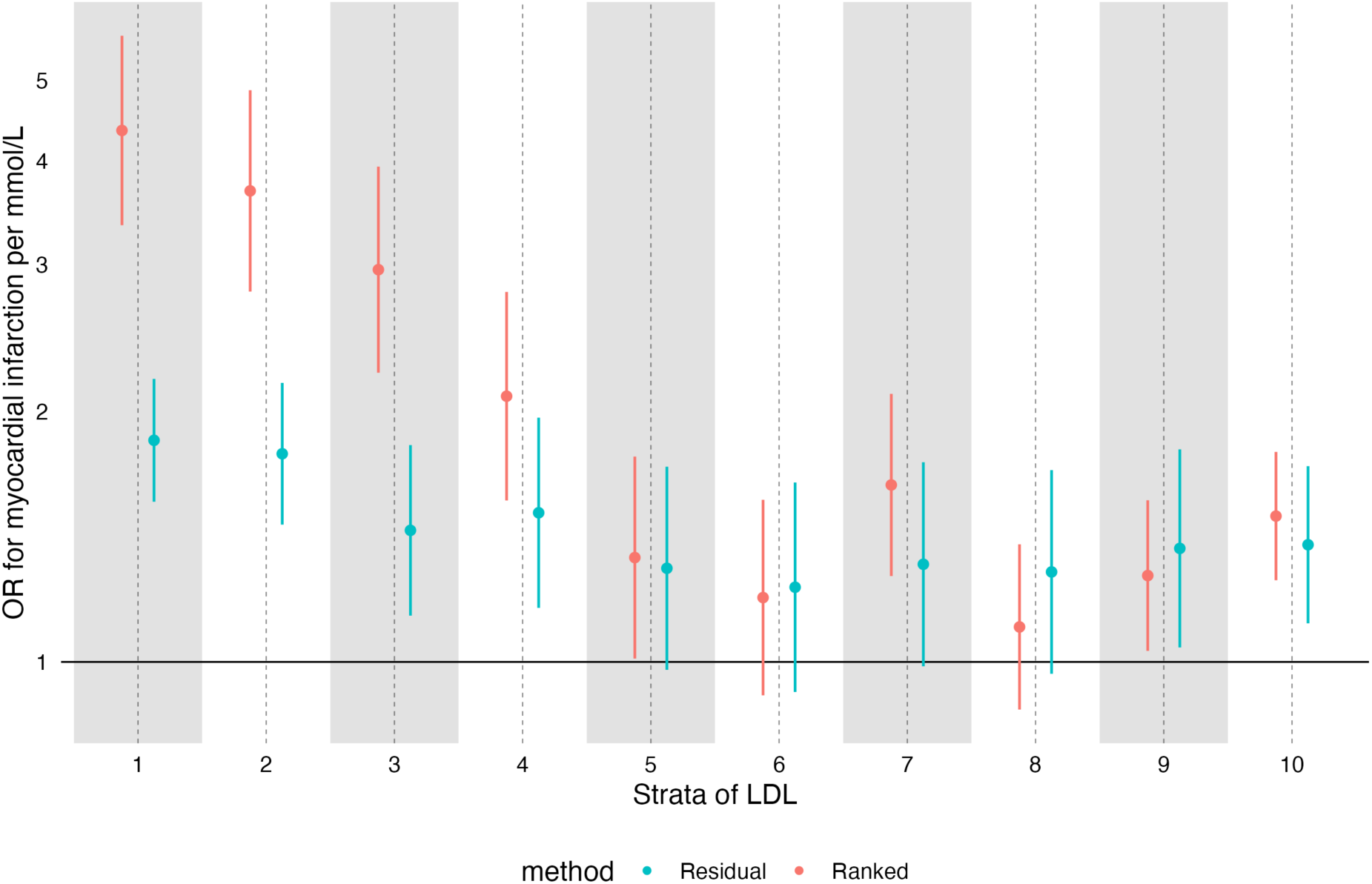
The estimated causal effect of increased LDL-C on MI. Estimates were generated in each stratum using the residual method (blue) and the doubly-ranked method (red) and were unadjusted for covariates.

For the residual method, the effect of LDL-C on MI was broadly stable and positive (although the trend still suggested reduction across strata, p = 0.02). When running analyses adjusting for age, sex, and the first 5 principal components, effects were similar but less extreme for MI (**Supplementary Figure S5)**, although there was still a clear negative trend (p = 0.001 for using the doubly-ranked method), with effect estimates much higher in those with lower LDL-C than those with higher LDL-C.

We ran sensitivity analyses for LDL-C on MI that included a) adjusting for statin use as a covariate b) in statin users and non-statin users, and c) in under 50s, where statin use was rare - 5.1% (**Supplementary Figure S6)**. We recognise these estimates adjusting for statin use are highly likely to be biased due to collider bias but include these for interest. As expected, stratifying or adjusting for statin usage altered estimates dramatically. When adding statin as a covariate, estimates of LDL-C favoured protection in upper and lower strata, but were null in the middle strata (u-shaped). When analyses were performed using those only under 50, estimates seemed similar to our primary analyses but showed more variability. Similarly, analyses restricted to non-statin users looked similar to our primary analyses. Analyses in statin users had reversed estimates in most strata, with increased LDL-C associated with reduced risk of MI.

### Triglycerides and cancer mortality

In the recent pre-print on non-linear effects of lipids[29], one analysis focussed on the effect of triglycerides on cancer mortality. The overall effect was null in both univariable and multivariable MR, but the authors report extremely implausible results: a strong positive effect in strata 1 (of ten): an OR of 2.57 (95% CI 1.67 to 3.96); but then a strong negative effect in strata 2: OR 0.56 (95% CI 0.39 to 0.83). All other strata specific estimates are close to the null. We report these estimates in **Figure 5**. A Z-test comparing the lower two strata has a p value of 1.4 x 10^-7^, while the overall p-value for heterogeneity is 9 x 10^-4^. These results are especially implausible given the known variability in measurements of triglycerides [46,47]. Therefore we aimed to replicate this as closely as possible, using the same dataset, exposure, outcome, and covariates. In our analysis (**Figure 6)**, we were unable to replicate this finding, despite the correlation between our strata specific mean triglyceride levels being >0.99, suggesting we are analysing the same strata.

**Figure 5:**
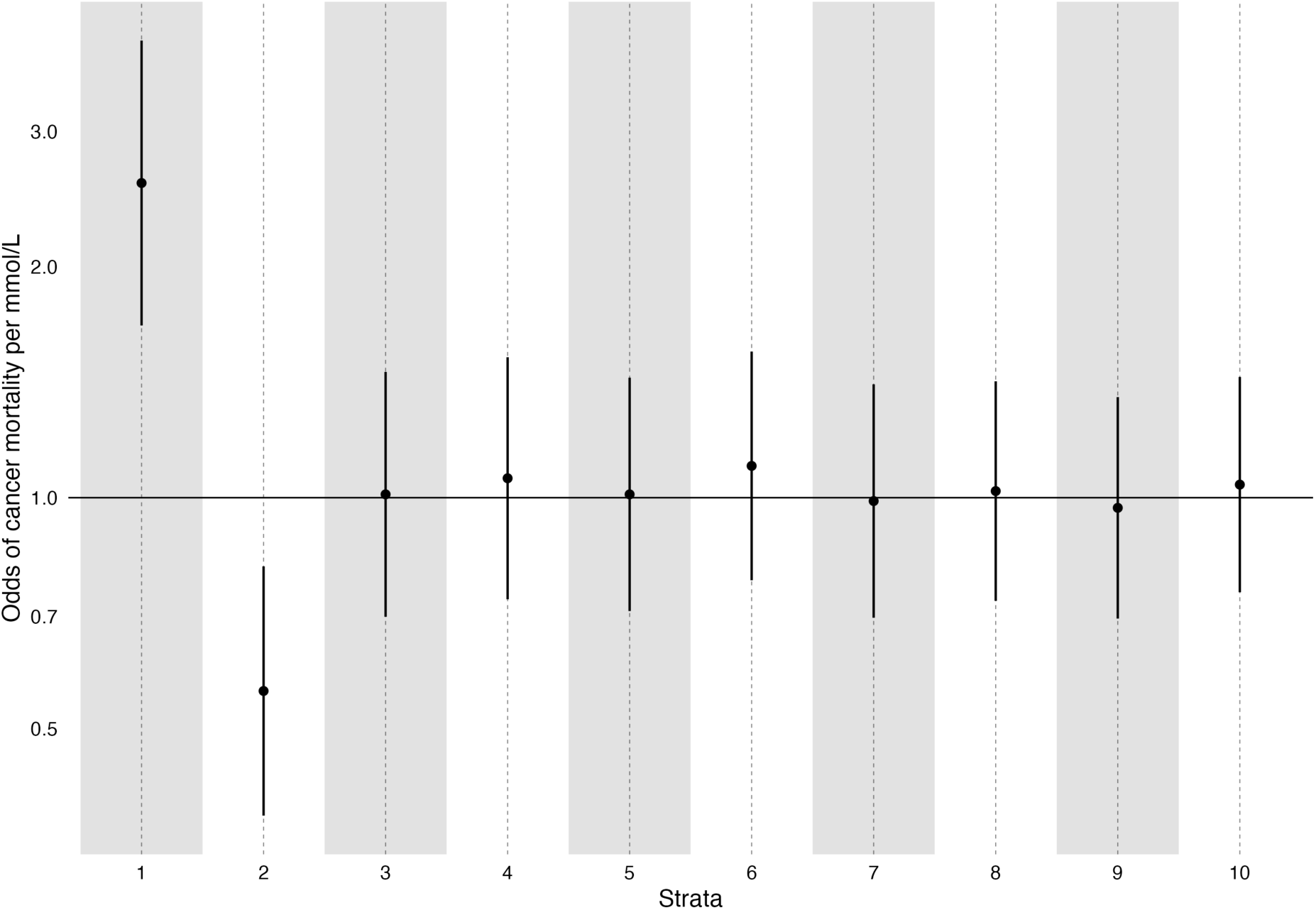
Estimates of the effect of triglycerides on cancer mortality from Yang et al [29]. Strata specific estimates generated using the doubly-ranked method.[29]

**Figure 6:**
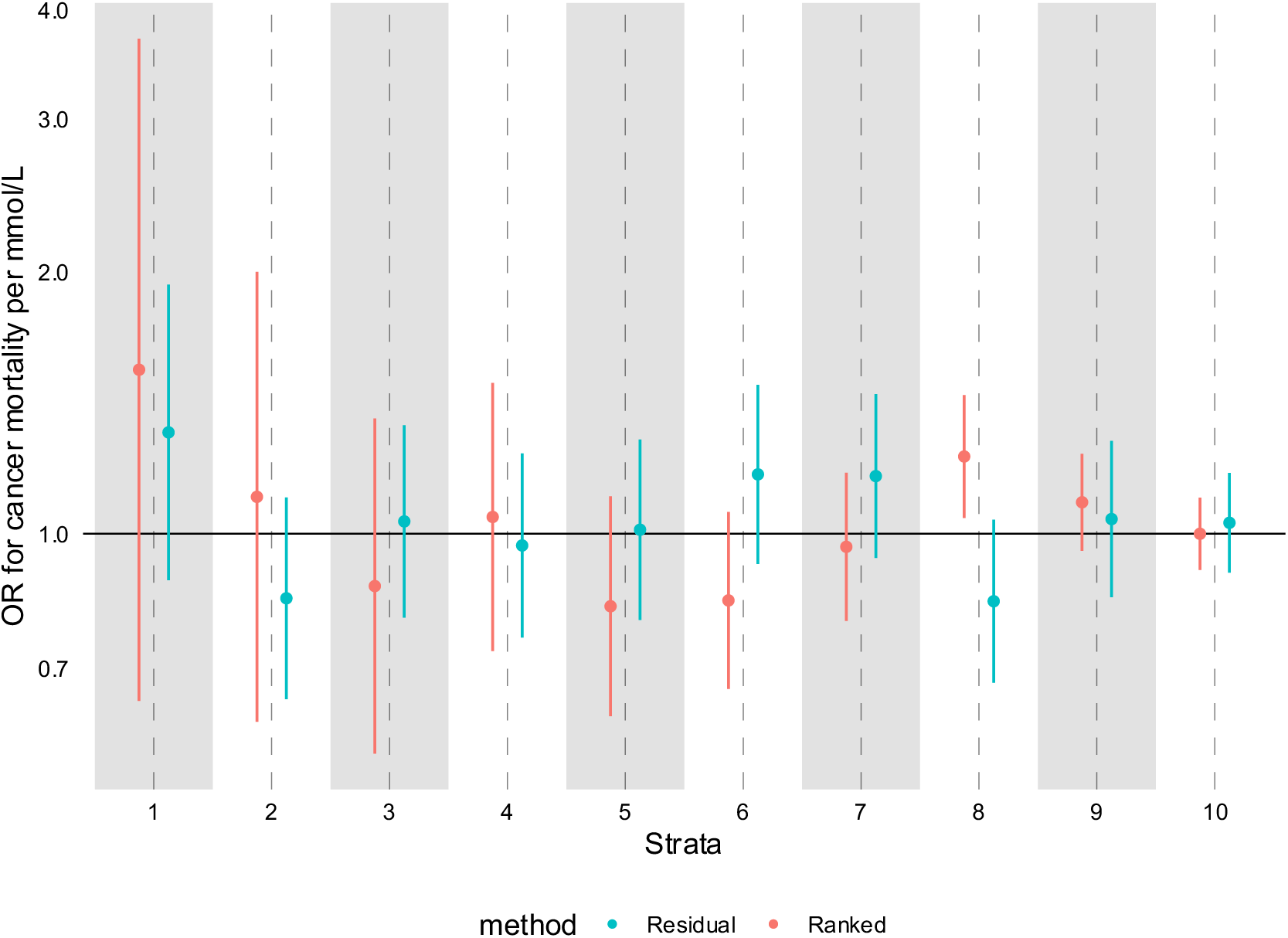
Estimates of the effect of triglycerides on cancer mortality using both the doubly-ranked (red) and residual method (blue). Estimates were generated adjusting for age, sex, age * sex, age * age, age * age * sex, and the first ten principle components.

To further assess whether the prior analysis was unreliable, we used the repeat sampling data from UK Biobank, that was taken approximately 2 years after the original visit. Data on triglyceride levels at both time points was available for 13,535 participants. Correlation between each time point was moderate (Pearson’s R 0.60). As expected, when generating strata using the doubly-ranked method on the original and repeat sample, participants were often not classified in the same strata. In fact, only 37% of those participants originally in strata 1 remain in strata 1, with 22% of them in strata 2, and the rest in higher strata. For those participants originally in strata 2, only 20% remain in strata 2, with 22% now in strata 1, and the rest in higher strata. These results are visualised in the alluvial plot below (**Figure 7)**.

**Figure 7:**
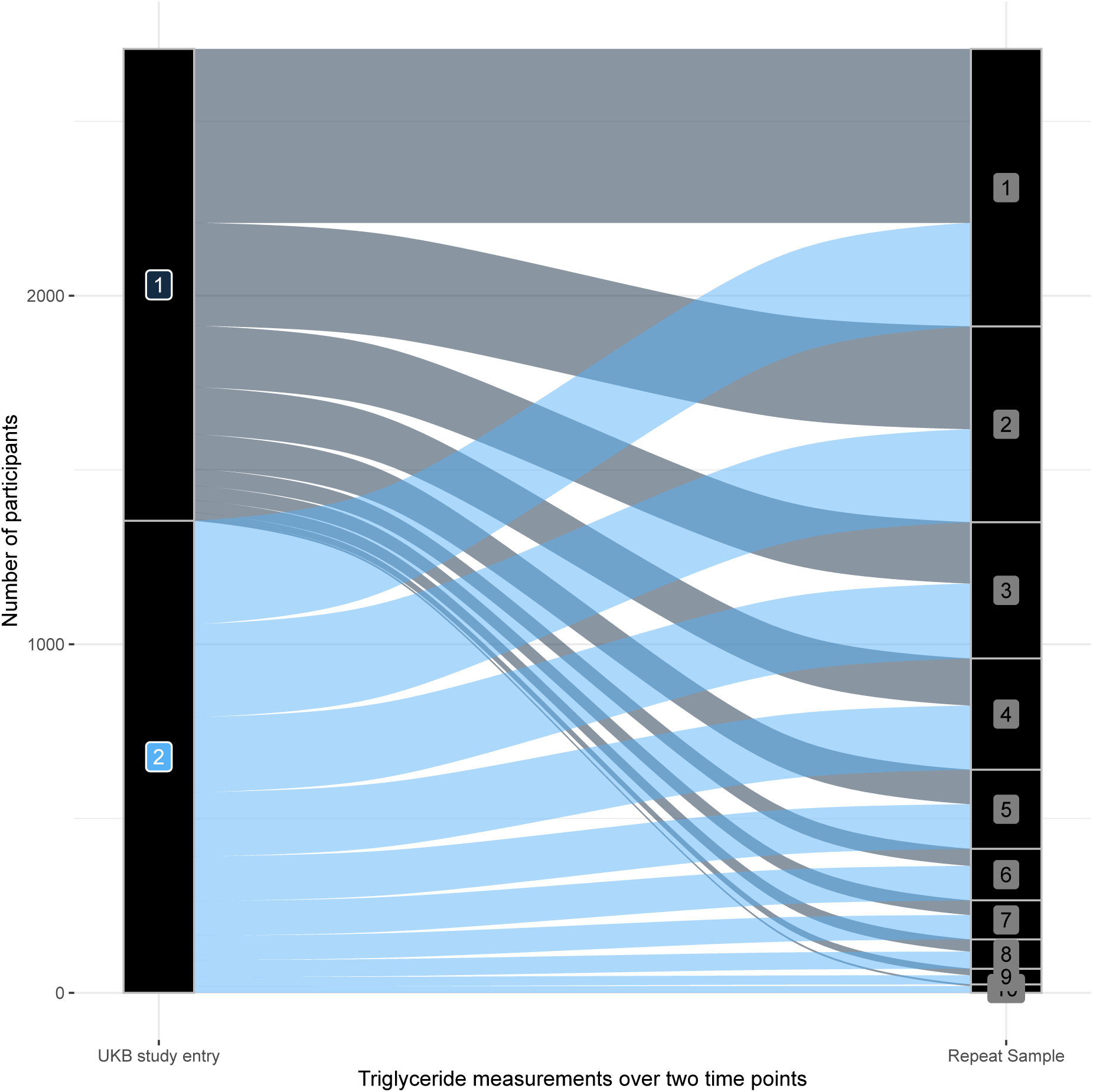
Classification of 13,535 participants who have repeat measurements into doubly-ranked strata based on triglyceride levels. The left hand Y axis represents the original strata, the right hand Y-axis the strata at repeat sampling.

This variability is not consistent with the reported estimates from Yang et al, which would suggest those in strata 1 are at greatly increased risk of cancer mortality, and those in strata 2 at greatly reduced risk, with no effect elsewhere. If this were the case, more than half of the participants in the lower two strata dramatically change their risk of cancer mortality over 2 years, with some estimates flipping from greatly increased risk to protective. To present analyses which implicitly assumes this is possible is simply not credible..

## Discussion

Mendelian randomization has become a commonly used approach in biomedical research[3], with the original intention being to strengthen causal inference regarding the effect of modifiable exposures on health outcomes [1]. Quantified estimates of various average benefits that would be seen from treatment are often presented in MR papers [2], with the more recent development of methods that allowed for estimation of the effect of such treatments in groups at differing levels of the exposure[7–10,48]. The residual method [10] is currently the most widely used approach. However recent findings have identified that this method can be seriously biased [19] leading to estimates that are inconsistent with causal interpretation. In particular, the central estimate for some outcomes was precisely estimated and close to the null, while all strata specific estimates favoured protection. The authors of this paper and the residual method accept this is not possible and referred to it “as a logical impossibility if all estimates have a causal interpretation”[23] after the journal retracted the paper[24] following being alerted to it being obvious seriously problematic[19]. However, whether the recently developed doubly-ranked method also leads to biased results remains unknown, leading us to perform our negative control outcome analyses.

In our negative control outcome analyses we identified that both methods showed bias, with effect estimates on age and sex differing across generated strata, when estimates should be null across all strata. It is worth considering some of the potential reasons for the distorted estimates in our negative control outcomes. One explanation is that these relate to a form of selection bias into strata which lead to associations between the genetic variants used to proxy for the exposure in the MR analyses – the so-called “instruments” – and other factors, including age and sex. This essentially reintroduces bias in MR analyses that were advanced with the intention of producing unbiased evidence on effects of exposures[1,2].

For body mass index overall selection bias (on entering studies, not on entering strata) is known to distort estimates, and thus the non-linear analysis of this exposure may be particularly liable to bias. For example, a large (∼ 3.3 million person) GWAS of chromosomal sex identified over 150 autosomal loci associated with male sex, including the body mass increasing allele at *FTO* (OR 1.02, *P* = 4.4 × 10^-36^) [40]. Our findings may in part reflect the strong associations between age, sex, and body mass index in the selected population of UK Biobank. This would be supported by the weaker association with Vitamin D. However, this explanation is potentially less appealing for the effect of LDL-C – with the most extreme non-null estimates. Although there is likely selection onto LDL-C, there is little evidence that adjusting for participant bias in UK Biobank affects MR estimates in LDL-C [49].

Additionally, these NCO results could represent population stratification differing across strata, which can be uncovered using negative controls [26]. Thirdly, these could represent methods related issues that occur on generation of the strata unrelated to selection. As we have demonstrated, stratification by both methods can lead to substantial alteration of estimates of IV-exposure associations. Regardless of the reason for the bias, these findings support more widespread use of negative control outcomes in Mendelian randomization studies, alongside a closer look at model assumptions.

Our analysis on the effect of LDL-C on myocardial infarction raises further concerns. The evidence base supporting the effectiveness of LDL-C reduction for MI is among the strongest in modern medicine, with >30 randomised trials to date showing benefit [27]. As such, meta-regressions of these trials using their baseline LDL-C have confidently suggested increased benefit of LDL-C reduction in those with higher LDL-C. This is also consistent with decades of non-RCT evidence, and our understanding of the pathogenesis of atherosclerosis (see **Box 1** for further background). In non-linear MR (especially with the doubly-ranked method), we identified the *opposite* non-linear effect, suggesting a *decreased* effectiveness of LDL-C reduction in those with higher LDL-C. Importantly, this bias was not ameliorated by the addition of age, sex, and genetic principal components into the model. Estimates were also similar whether using incident, prevalent, or other MI, so these results are not likely to be generated by survivorship bias.

In fact, these results do not depend on our particular analytical approach and/or outcome definition. A recent preprint from the authors of the doubly-ranked method performed a similar analysis on the same dataset [29] (using coronary artery disease as an outcome, not MI), and found similar findings, and with a strong negative trend with increasing LDL-C reduction (**Figure 8** compares both analyses). This consistent disagreement between trial data and MR data suggests a fundamental issue with the method, or with the application of this method to this data. This may reflect selection, but importantly, estimates were broadly similar in our primary analysis (unadjusted for age, sex and genetic principal components) and their analysis (which adjusts for the above), and suggests that if selection bias is occurring, it is not fixed by adjusting for age and sex.

**Figure 8:**
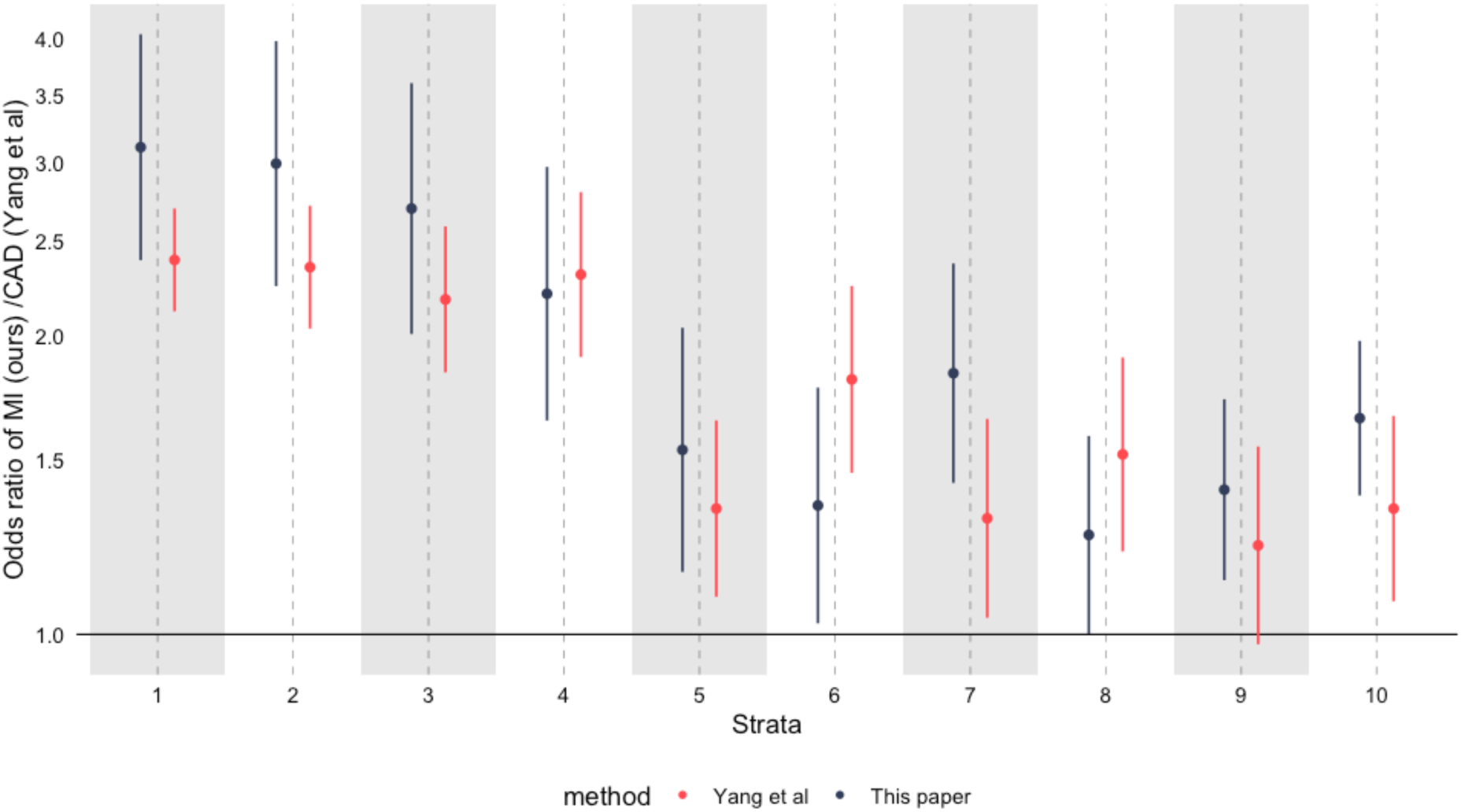
The effect of increasing LDL-C on MI (our data), and coronary artery disease (CAD, Yang et al[29]). Both estimates adjusted for age, sex, and genetic principal components and using UK Biobank participants. Red estimates from Yang et al. Both estimates using the doubly-ranked method.

It is therefore plausible these findings relate to other issues such as statin use (which has a large effect on LDL-C). However, adjusting for statins did not recover sensible estimates, nor would this be advised as this would lead to collider bias. Even if sensible estimates were recovered, this would suggest that non-linear MR estimates require understanding and modelling of the covariate structure of the exposure and the outcome, which would bring in all the issues of traditional observational analyses that MR is designed to remove[1,2]. This particular example of LDL-C on CAD is particularly challenging for non-linear MR, as the strength of evidence supporting the effectiveness of conventional MR for the effect of LDL-C on CAD is very high, with multiple drugs having strong support from MR analyses [50].

Further implausible analyses have been generated in the same preprint with an analysis on TG and cancer mortality. It is not possible to think of a plausible biological model where those in the bottom strata have very robustly estimated *increased* cancer mortality with higher triglycerides, whilst shifting into the 2^nd^ lowest lowest strata have robustly estimated *decreased* cancer mortality, with there being no causal effect for all other participants. In addition given the considerable fluctuations demonstrated in triglyceride levels individuals would shift their classification in one into the other strata depending on exactly when the measurement for the particular individuals were made. Importantly, we were unable to replicate the findings in this preprint[29] despite being able to generate strata specific mean estimates of TG that exactly matched theirs, and using extremely similar definitions and analytical approaches, and despite being able to replicate similar findings for the effect of LDL-C on MI as they report on CAD.

Therefore, our findings – and the results of other, independently performed analyses - suggest that we should be cautious when applying either the residual or doubly-ranked MR methods, as both methods seem potentially more susceptible to bias in the generation of strata specific estimates than conventional MR estimates are. Whether this entirely relates to selection bias or to other methodological issues remains unknown. Corrections should be issued for papers which have produced likely misleading estimates of non-linear effects [20,21,24], especially when their misleading findings gained widespread publicity due to the efforts of authors [51]. That these biases relate only to the examples of body mass index, Vitamin D, and lipid traits is unlikely, but they remain to be demonstrated for other exposures.

### Limitations

This paper provides an initial assessment of the doubly doubly-ranked and residual methods for performing non-linear MR. In particular, our analysis focussed on three exposures (BMI, Vitamin D and lipids) a small number of outcomes and included only one dataset (UK Biobank), which we limited to White British participants. It is possible that our findings do not extend to all exposures, or only occur in certain settings. However, nearly all published non-linear analyses use UK Biobank, so these limitations likely apply to much of the published literature. Further analyses should examine these methods on diverse datasets, exposures, and outcomes, with proof of principle examples.

## Conclusion

Using a negative control outcome approach, we identified bias in non-linear mendelian randomization estimates of the effect of BMI, Vitamin D and LDL-C using two commonly used methods. Estimates of the effect of LDL-C on cardiovascular outcomes were inconsistent with RCTs and biological understanding. Until reliable evidence is presented that the methods are generating sensible findings there should be a moratorium on the further publication of non-linear MR findings using the two methods we examine in this paper.

## Data Availability

This analysis was performed in the UK Biobank. Data can be accessed via applicaton to the UK Biobank. We provide code used to perform our analysis in a supplement.

## Statements and Declarations

### Author contributions

GDS conceived the idea, while FH performed analyses and wrote the initial manuscript. DH, WS, and KT provided commentary and feedback.

## References

1. Davey Smith G, Ebrahim S. ‘Mendelian randomization’: can genetic epidemiology contribute to understanding environmental determinants of disease?*. Int J Epidemiol [Internet]. 2003 [cited 2023 May 22];32:1–22. Available from: https://academic.oup.com/ije/article/32/1/1/642797

2. Sanderson E, Glymour MM, Holmes MV, Kang H, Morrison J, Munafò MR, et al. Mendelian randomization. Nature Reviews Methods Primers [Internet]. 2022 [cited 2023 May 22];2:1–21. Available from: https://www.nature.com/articles/s43586-021-00092-5

3. Smith GD, Ebrahim S. Mendelian randomisation at 20 years: how can it avoid hubris, while achieving more? Lancet Diabetes Endocrinol [Internet]. 2024;12:14–7. Available from: 10.1016/S2213-8587(23)00348-0

4. Sobczyk MK, Zheng J, Davey Smith G, Gaunt TR. Systematic comparison of Mendelian randomisation studies and randomised controlled trials using electronic databases. BMJ Open [Internet]. 2023;13:e072087. Available from: 10.1136/bmjopen-2023-072087

5. Lawlor DA, Tilling K, Davey Smith G. Triangulation in aetiological epidemiology. Int J Epidemiol [Internet]. 2016;45:1866–86. Available from: 10.1093/ije/dyw314

6. Munafò MR, Higgins JPT, Davey Smith G. Triangulating Evidence through the Inclusion of Genetically Informed Designs. Cold Spring Harb Perspect Med [Internet]. 2021;11. Available from: 10.1101/cshperspect.a040659

7. Staley JR, Burgess S. Semiparametric methods for estimation of a nonlinear exposure-outcome relationship using instrumental variables with application to Mendelian randomization. Genet Epidemiol [Internet]. 2017;41:341–52. Available from: 10.1002/gepi.22041

8. Sulc J, Sjaarda J, Kutalik Z. Polynomial Mendelian randomization reveals non-linear causal e]ects for obesity-related traits. HGG Adv [Internet]. 2022;3:100124. Available from: 10.1016/j.xhgg.2022.100124

9. Silverwood RJ, Holmes MV, Dale CE, Lawlor DA, Whittaker JC, Davey Smith G, et al. Testing for non-linear causal effects using a binary genotype in a Mendelian randomization study: application to alcohol and cardiovascular traits. Int J Epidemiol [Internet]. 2014;43:1781–90. Available from: 10.1093/ije/dyu187

10. Burgess S, Davies NM, Thompson SG, EPIC-InterAct Consortium. Instrumental variable analysis with a nonlinear exposure-outcome relationship. Epidemiology [Internet]. 2014;25:877–85. Available from: 10.1097/EDE.0000000000000161

11. Tian H, Mason AM, Liu C, Burgess S. Relaxing parametric assumptions for non-linear Mendelian randomization using a doubly-ranked stratification method. PLoS Genet [Internet]. 2023 [cited 2023 May 22];19:e1010823. Available from: 10.1371/journal.pgen.1010823

12. Rogne T, Solligård E, Burgess S, Brumpton BM, Paulsen J, Prescott HC, et al. Body mass index and risk of dying from a bloodstream infection: A Mendelian randomization study. PLoS Med [Internet]. 2020;17:e1003413. Available from: 10.1371/journal.pmed.1003413

13. Pinto Pereira SM, Garfield V, Norris T, Burgess S, Williams DM, Dodds R, et al. Linear and Non-linear associations between vitamin D and grip strength: a Mendelian Randomisation study in UK Biobank. J Gerontol A Biol Sci Med Sci [Internet]. 2022; Available from: 10.1093/gerona/glac255

14. Zhou A, Hyppönen E. Vitamin D deficiency and C-reactive protein: a bidirectional Mendelian randomization study. Int J Epidemiol [Internet]. 2023;52:260–71. Available from: 10.1093/ije/dyac087

15. Sutherland JP, Zhou A, Hyppönen E. Vitamin D Deficiency Increases Mortality Risk in the UK Biobank : A Nonlinear Mendelian Randomization Study. Ann Intern Med [Internet]. 2022;175:1552–9. Available from: 10.7326/M21-3324

16. Zhou A, Selvanayagam JB, Hyppönen E. Non-linear Mendelian randomization analyses support a role for vitamin D deficiency in cardiovascular disease risk. Eur Heart J [Internet]. 2022;43:1731–9. Available from: 10.1093/eurheartj/ehab809

17. Sun Y-Q, Burgess S, Staley JR, Wood AM, Bell S, Kaptoge SK, et al. Body mass index and all cause mortality in HUNT and UK Biobank studies: linear and non-linear mendelian randomisation analyses. BMJ [Internet]. 2019;364:l1042. Available from: 10.1136/bmj.l1042

18. Sofianopoulou E, Kaptoge SK, Afzal S, Jiang T, Gill D, Gundersen TE, et al. RETRACTED: Estimating dose-response relationships for vitamin D with coronary heart disease, stroke, and all-cause mortality: observational and Mendelian randomisation analyses. The Lancet Diabetes & Endocrinology [Internet]. 2021;9:837–46. Available from: https://www.sciencedirect.com/science/article/pii/S2213858721002631

19. Davey Smith G. Mendelian randomisation and vitamin D: the importance of model assumptions [Internet]. Lancet Diabetes Endocrinol. Elsevier BV; 2023. p. 14. Available from: 10.1016/S2213-8587(22)00345-X

20. Burgess S, Butterworth AS. Dose-response relationships for vitamin D and all-cause mortality - Authors’ reply [Internet]. Lancet Diabetes Endocrinol. 2022. p. 158–9. Available from: 10.1016/S2213-8587(22)00015-8

21. Wade KH, Hamilton FW, Carslake D, Sattar N, Davey Smith G, Timpson NJ. Challenges in undertaking non-linear Mendelian randomization. Obesity [Internet]. 2023;2887–90. Available from: 10.1002/oby.23927

22. Munafò MR, Tilling K, Taylor AE, Evans DM, Davey Smith G. Collider scope: when selection bias can substantially influence observed associations. Int J Epidemiol [Internet]. 2018;47:226–35. Available from: 10.1093/ije/dyx206

23. Emerging Risk Factors Collaboration/EPIC-CVD/Vitamin D Studies Collaboration. Estimating dose-response relationships for vitamin D with coronary heart disease, stroke, and all-cause mortality: observational and Mendelian randomisation analyses. Lancet Diabetes Endocrinol. 2024;9:837–46.

24. The Editors Of The Lancet Diabetes Endocrinology. Retraction and republication-Estimating dose-response relationships for vitamin D with coronary heart disease, stroke, and all-cause mortality: observational and Mendelian randomisation analyses. Lancet Diabetes Endocrinol [Internet]. 2023; Available from: 10.1016/S2213-8587(23)00364-9

25. Burgess S. Violation of the constant genetic effect assumption can result in biased estimates for non-linear Mendelian randomization [Internet]. Hum. Hered. 2023 [cited 2022 Nov 1]. p. 2022.10.26.22280570. Available from: 10.1159/000531659

26. Sanderson E, Richardson TG, Hemani G, Davey Smith G. The use of negative control outcomes in Mendelian randomization to detect potential population stratification. Int J Epidemiol [Internet]. 2021;50:1350–61. Available from: 10.1093/ije/dyaa288

27. Navarese EP, Robinson JG, Kowalewski M, Kolodziejczak M, Andreotti F, Bliden K, et al. Association Between Baseline LDL-C Level and Total and Cardiovascular Mortality After LDL-C Lowering: A Systematic Review and Meta-analysis. JAMA [Internet]. 2018;319:1566–79. Available from: 10.1001/jama.2018.2525

28. Cholesterol Treatment Trialists’ (CTT) Collaboration. Efficacy and safety of more intensive lowering of LDL cholesterol: a meta-analysis of data from 170 000 participants in 26 randomised trials. Lancet [Internet]. 2010;376:1670–81. Available from: 10.1016/S0140-6736(10)61350-5

29. Yang G, Mason AM, Wood AM, Schooling CM, Burgess S. Assessing dose-response relations of lipid traits with coronary artery disease, all-cause mortality, and cause-specific mortality: a linear and non-linear Mendelian randomization study [Internet]. bioRxiv. 2023. Available from: https://www.medrxiv.org/content/10.1101/2023.09.27.23296203v1.abstract

30. Bycroft C, Freeman C, Petkova D, Band G, Elliott LT, Sharp K, et al. The UK Biobank resource with deep phenotyping and genomic data. Nature [Internet]. 2018;562:203–9. Available from: 10.1038/s41586-018-0579-z

31. Mitchell R, Hemani G, Dudding T, Corbin L, Harrison S, Paternoster L. UK Biobank genetic data: MRC-IEU quality control, version 2 [Internet]. University of Bristol; 2019 [cited 2021 Dec 21]. Available from: https://data.bris.ac.uk/data/dataset/1ovaau5sxunp2cv8rcy88688v

32. Liu H, Li J, Liu F, Huang K, Cao J, Chen S, et al. Efficacy and safety of low levels of low-density lipoprotein cholesterol: trans-ancestry linear and non-linear Mendelian randomization analyses. Eur J Prev Cardiol [Internet]. 2023;30:1207–15. Available from: 10.1093/eurjpc/zwad111

33. Locke AE, Kahali B, Berndt SI, Justice AE, Pers TH, Day FR, et al. Genetic studies of body mass index yield new insights for obesity biology. Nature [Internet]. 2015 [cited 2022 Apr 28];518:197–206. Available from: 10.1038/nature14177

34. Hemani G, Zheng J, Elsworth B, Wade KH, Haberland V, Baird D, et al. The MR-Base platform supports systematic causal inference across the human phenome. Elife [Internet]. 2018;7. Available from: 10.7554/eLife.34408

35. Wang TJ, Zhang F, Richards JB, Kestenbaum B, van Meurs JB, Berry D, et al. Common genetic determinants of vitamin D insufficiency: a genome-wide association study [Internet]. Lancet. 2010. p. 180–8. Available from: 10.1016/S0140-6736(10)60588-0

36. Purcell S, Neale B, Todd-Brown K, Thomas L, Ferreira MAR, Bender D, et al. PLINK: a tool set for whole-genome association and population-based linkage analyses. Am J Hum Genet [Internet]. 2007;81:559–75. Available from: 10.1086/519795

37. Choi Y, Chan AP, Kirkness E, Telenti A, Schork NJ. Comparison of phasing strategies for whole human genomes. PLoS Genet [Internet]. 2018;14:e1007308. Available from: 10.1371/journal.pgen.1007308

38. Howie B, Marchini J, Stephens M. Genotype imputation with thousands of genomes. G3 [Internet]. 2011;1:457–70. Available from: 10.1534/g3.111.001198

39. Sanderson E, Macdonald-Wallis C, Davey Smith G. Negative control exposure studies in the presence of measurement error: implications for attempted effect estimate calibration. Int J Epidemiol [Internet]. 2018;47:587–96. Available from: 10.1093/ije/dyx213

40. Pirastu N, Cordioli M, Nandakumar P, Mignogna G, Abdellaoui A, Hollis B, et al. Genetic analyses identify widespread sex-differential participation bias. Nat Genet [Internet]. 2021;53:663–71. Available from: 10.1038/s41588-021-00846-7

41. Carreras-Torres R, Johansson M, Haycock PC, Relton CL, Davey Smith G, Brennan P, et al. Role of obesity in smoking behaviour: Mendelian randomisation study in UK Biobank. BMJ [Internet]. 2018 [cited 2023 Apr 26];361:k1767. Available from: 10.1136/bmj.k1767

42. Mason AM, Burgess S. Software Application Profile: SUMnlmr, an R package that facilitates flexible and reproducible non-linear Mendelian randomization analyses. Int J Epidemiol [Internet]. 2022 [cited 2023 May 22];51:2014–9. Available from: https://academic.oup.com/ije/article/51/6/2014/6659062

43. Palmer T, Spiller W, Sanderson E. OneSampleMR: One Sample Mendelian Randomization and Instrumental Variable Analyses. 2023.

44. Holmes MV, Davey Smith G. Dyslipidaemia: Revealing the effect of CETP inhibition in cardiovascular disease. Nat Rev Cardiol [Internet]. 2017;14:635–6. Available from: 10.1038/nrcardio.2017.156

45. Smith GD, Ebrahim S. Mendelian randomization: prospects, potentials, and limitations. Int J Epidemiol [Internet]. 2004 [cited 2024 Jan 18];33:30–42. Available from: https://academic.oup.com/ije/article-pdf/33/1/30/1939982/dyh132.pdf

46. Phillips AN, Smith GD. How independent are “independent” effects? Relative risk estimation when correlated exposures are measured imprecisely. J Clin Epidemiol [Internet]. 1991;44:1223–31. Available from: 10.1016/0895-4356(91)90155-3

47. Johansen MØ, Moreno-Vedia J, Balling M, Davey Smith G, Nordestgaard BG. Triglyceride content increases while cholesterol content decreases in HDL and LDL+IDL fractions following normal meals: The Copenhagen General Population Study of 25,656 individuals. Atherosclerosis [Internet]. 2023;383:117316. Available from: 10.1016/j.atherosclerosis.2023.117316

48. Millwood IY, Walters RG, Mei XW, Guo Y, Yang L, Bian Z, et al. Conventional and genetic evidence on alcohol and vascular disease aetiology: a prospective study of 500 000 men and women in China. Lancet [Internet]. 2019;393:1831–42. Available from: 10.1016/S0140-6736(18)31772-0

49. Schoeler T, Speed D, Porcu E, Pirastu N, Pingault J-B, Kutalik Z. Participation bias in the UK Biobank distorts genetic associations and downstream analyses. Nat Hum Behav [Internet]. 2023; Available from: 10.1038/s41562-023-01579-9

50. Allara E, Morani G, Carter P, Gkatzionis A, Zuber V, Foley CN, et al. Genetic Determinants of Lipids and Cardiovascular Disease Outcomes: A Wide-Angled Mendelian Randomization Investigation. Circ Genom Precis Med [Internet]. 2019;12:e002711. Available from: 10.1161/CIRCGEN.119.002711

51. The sunshine vitamin that ‘D’elivers on cardio health [Internet]. Home. [cited 2023 Sep 27]. Available from: https://www.unisa.edu.au/media-centre/Releases/2021/the-sunshine-vitamin-that-delivers-on-cardio-health/

