## Supplementary material for "Non-linear mendelian randomization: detection of biases using negative controls with a focus on BMI, Vitamin D and LDL cholesterol": Sup figures

**Figure S1:** The predicted causal effect of Vitamin D on age (A), and sex (B), and BMI on age (C) and sex (D). Estimates were generated in each stratum using the residual method (blue) and the doubly-ranked method (red) and were adjusted for age (for sex outcomes), for sex (for age outcomes), and the first 5 genetic principal components. The black estimate represents the conventional MR estimate for the whole cohort.

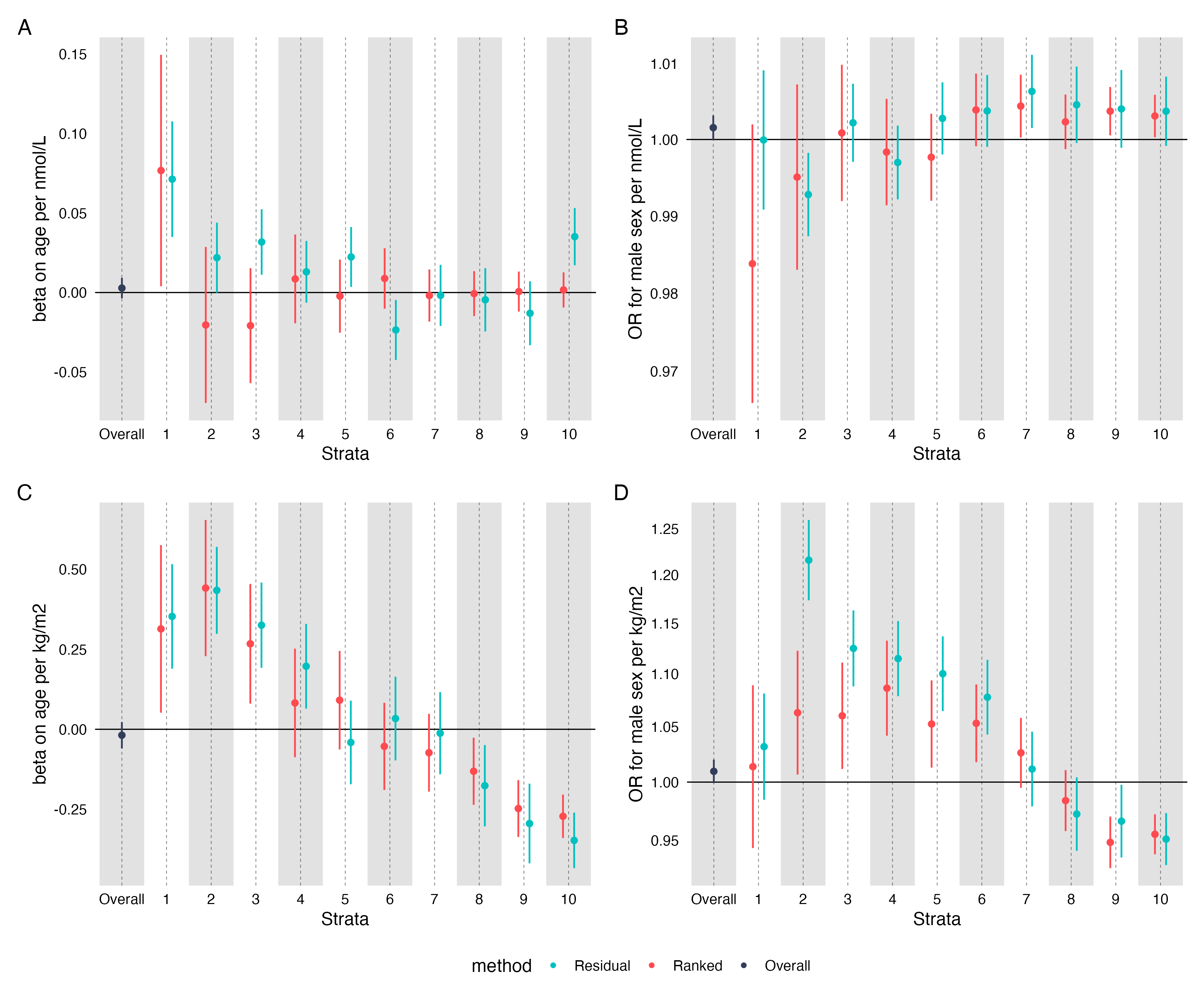

**Figure S2:** Strata specific MR associations between the residual and ranked model for the association between log transformed Vitamin D and age (A) and sex (B). Estimates were generated in each strata using the residual method (blue) and the doubly-ranked method (red).

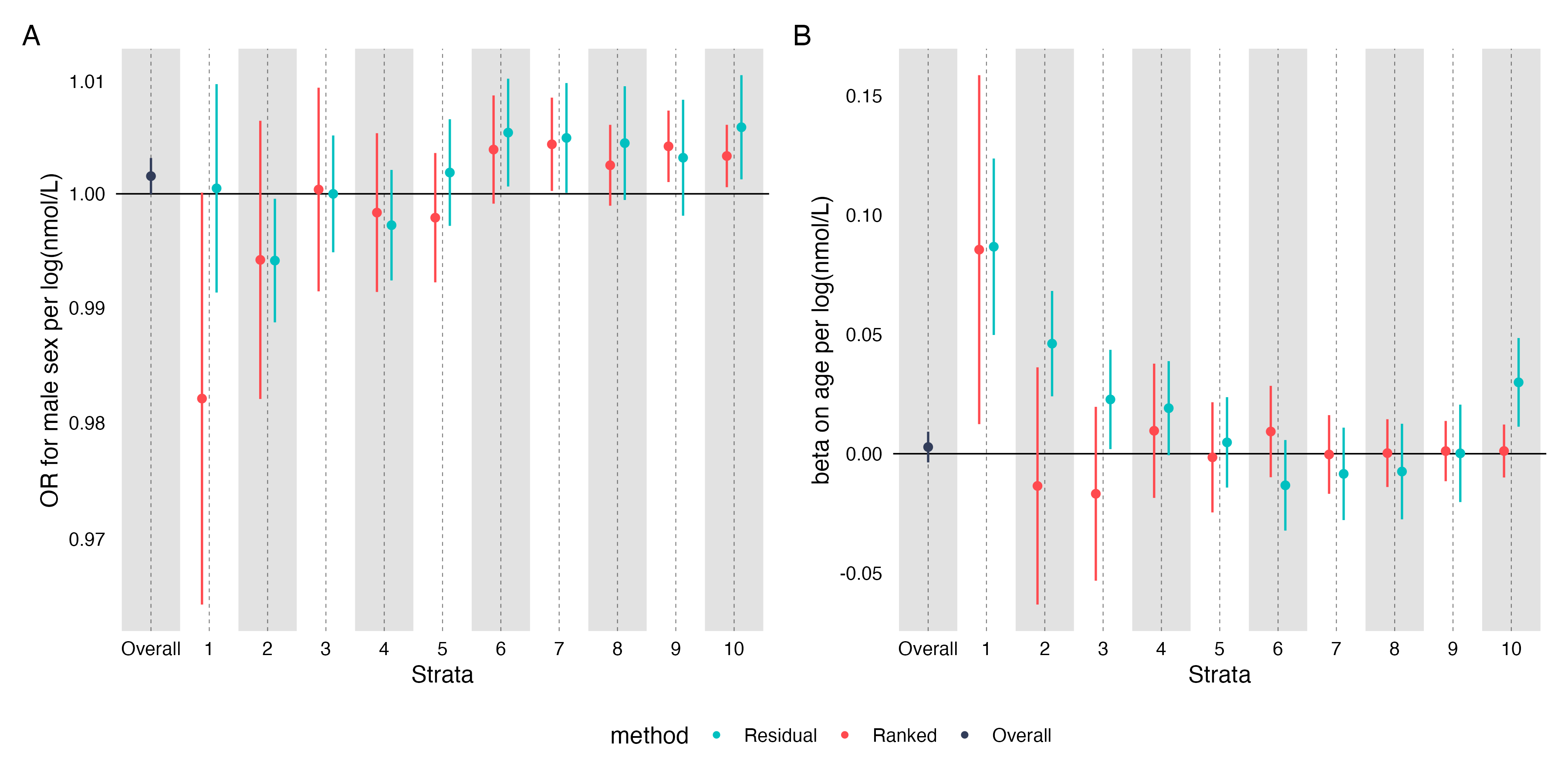

**Figure S3:** Strata specific MR estimates for the effect of BMI on sex (A) and age (B) in non-smokers, and on sex (C) and age(D) in smokers. Analysis performed for both the residual method (blue), and the doubly-ranked method (red). Conventional MR estimates for the sub cohort of smokers or non-smokers is in black.

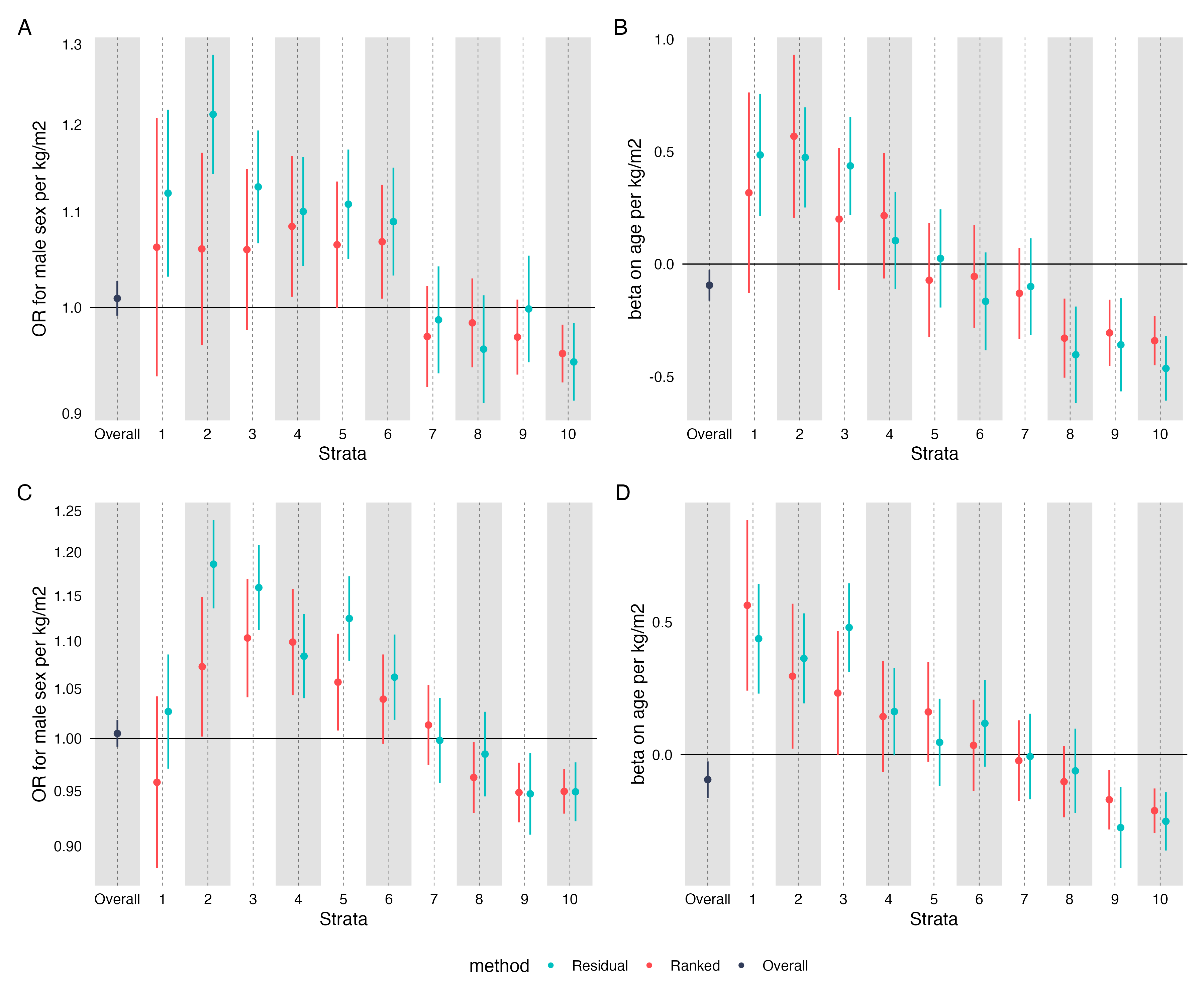

**Figure S4:** Non-linear MR estimates for the effect of LDL-C on prevalent MI (i.e. MI before recruitment to UK Biobank), and B) incident MI (i.e. MI after recruitment to UK Biobank). Analyses were run unadjusted for covariates. Estimates were generated in each stratum using the residual method (red) and the ranked method (black).

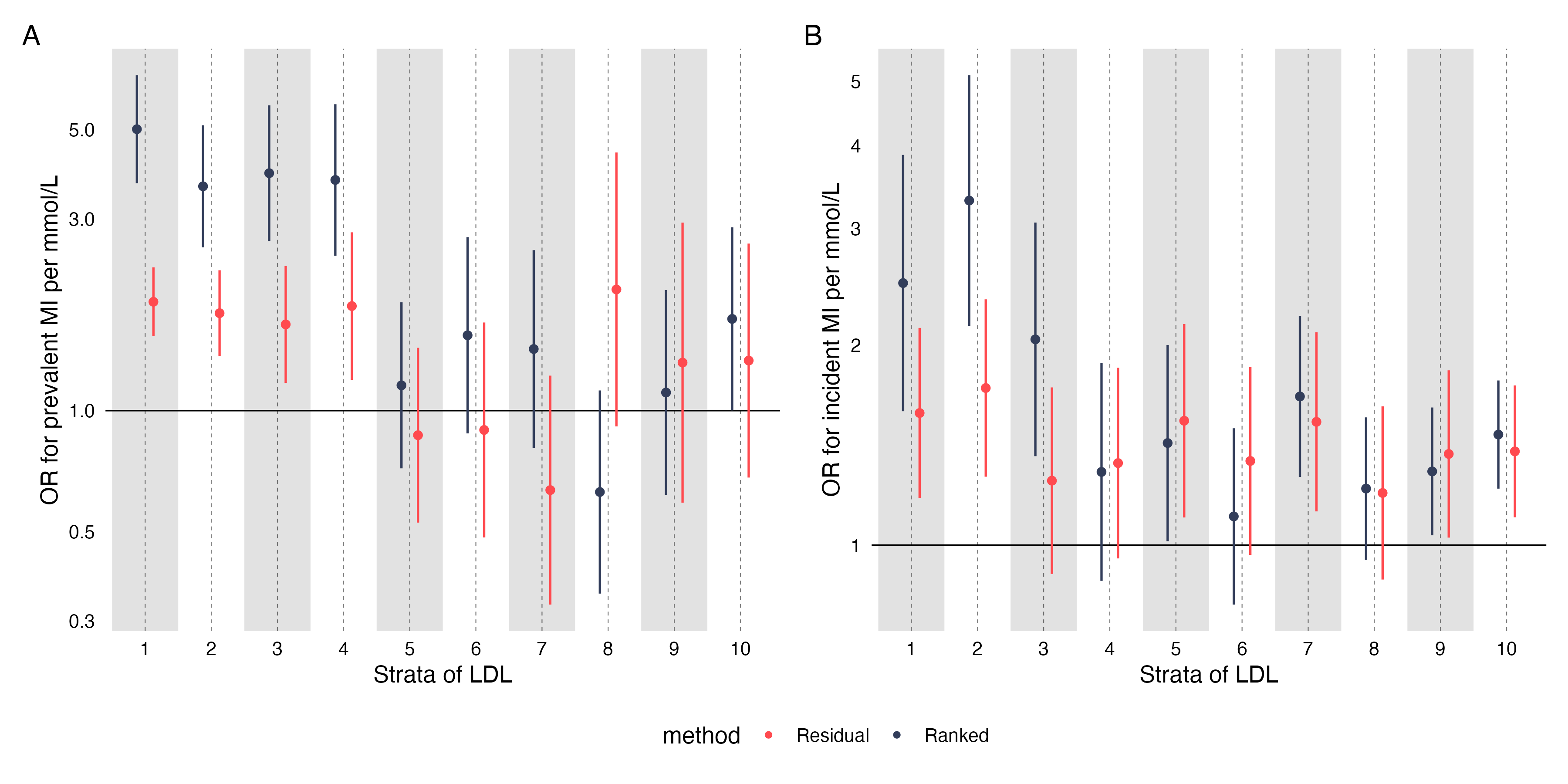

**Figure S5:** The predicted causal effect of increased LDL-C on MI. Estimates were generated in each stratum using the residual method (red) and the ranked method (black) and were adjusted for age, sex, and the first 5 genetic principal components.

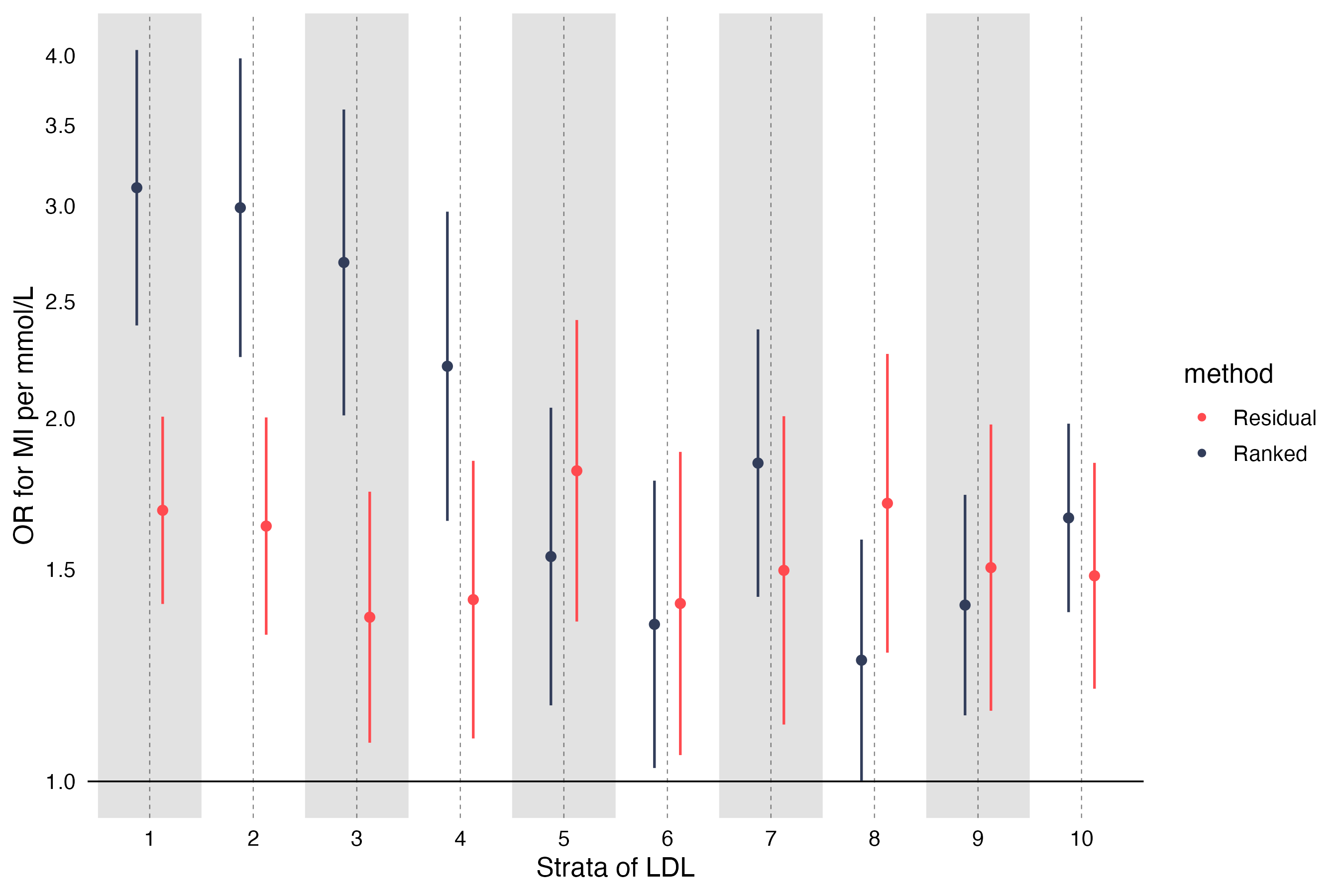

**Figure S6:** The predicted causal effect of increased LDL-C on MI in (A) analyses adjusting for statin users, sex, age, and the first 5 genetic principal components (B) UKB participants <50 years, (C), statin users, and (D), non-statin users. Estimates were generated in each stratum using the residual method (red) and the ranked method (black).

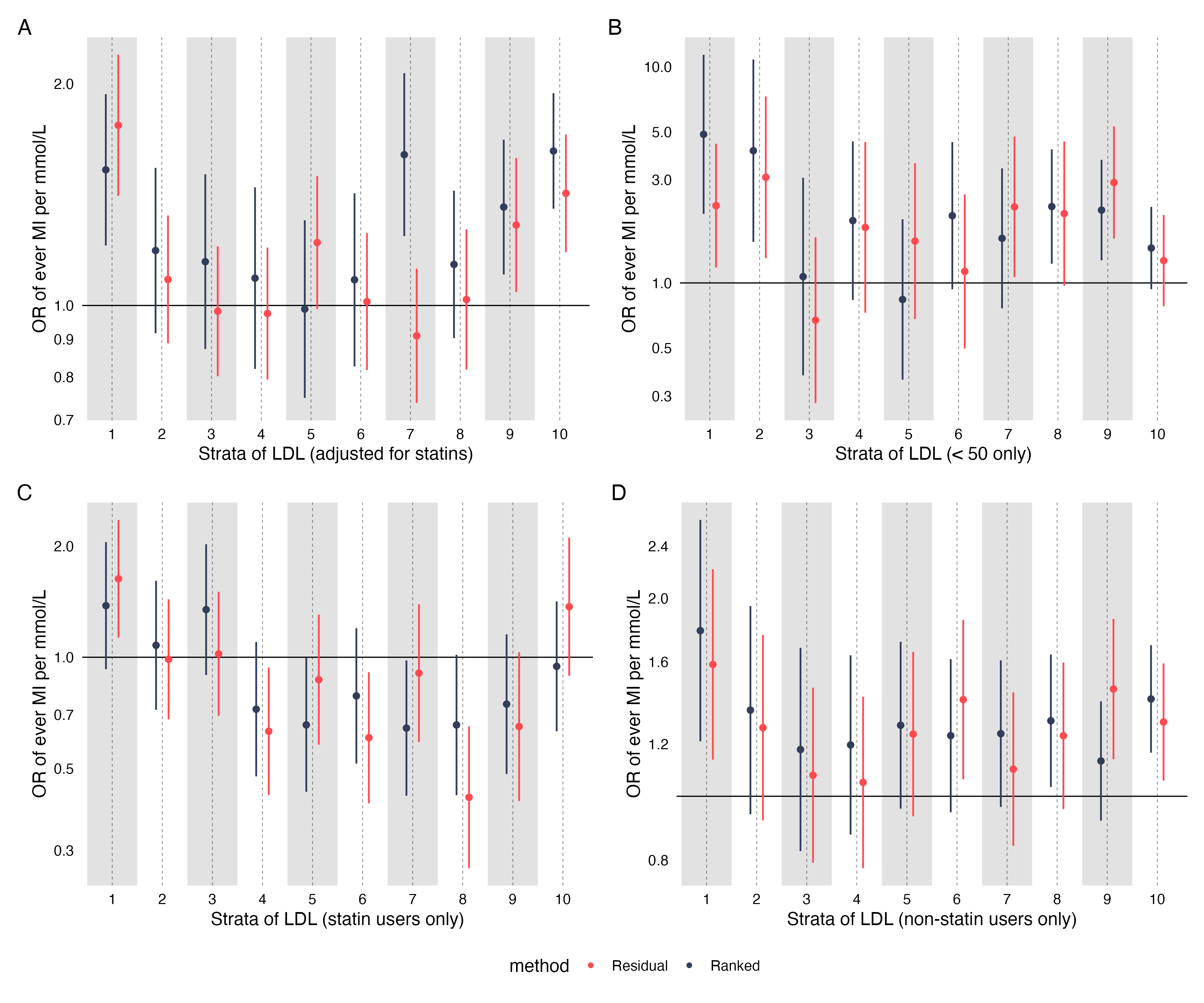
