## Supplementary material for "Non-linear mendelian randomization: detection of biases using negative controls with a focus on BMI, Vitamin D and LDL cholesterol": code: Non_linear_medrxiv_R1_LDL_EJE.qmd

````
---
title: "Analaysis of non-linear methods"
author: "Fergus Hamilton"
format: html
editor: visual
---

Medrxiv Non-linear piece.

```{r}
pacman::p_load(tidyverse, vroom, janitor, data.table, nlmr, SUMnlmr, broom, ggforestplot, patchwork, OneSampleMR)

```

TSLS function + strata generation

```{r}
generate_ranked_strata <- function(g, x, q, seed = "1") {
  z = rank(g, ties.method = "random")
  
  strata1 = floor((z-1)/q)+1
  
  # check GR statistic
  
  set.seed(seed)
  id= seq(x)
  temp<- data.frame(x=x,strata1=strata1,id=id, g=g)
  temp<- arrange(.data=temp, x)
  temp<-group_by(.data=temp, strata1)
  temp<-mutate(.data=temp, x0q= rank(x, ties.method = "random"))
  temp<-arrange(.data=temp, id)
  x0q <- temp$x0q
  return(x0q)
}
mr_using_prs_binary <- function(exposure, exposure_prs, outcome_list, data) {
  l <- list()
  for (i in 1:length(outcome_list)) {
    out <- as.data.frame(matrix(data = NA, nrow = 1))
    outcome <- outcome_list[i]
    print(outcome)
    tmp <- data %>% dplyr::select(all_of(c(exposure, exposure_prs, outcome_list)))
    tmp <- na.omit(tmp)
    attach(tmp)
    con = tmp %>% dplyr::filter((!!as.name(outcome)) == 0) %>% dplyr::select(all_of(exposure_prs)) %>% unlist #values for prs in the controls only
    lm <- lm(as.formula(paste0(exposure, '~', exposure_prs)),
             data = tmp %>% dplyr::filter((!!as.name(outcome)) == 0))
    predict.con = predict(lm, newdata=list(con=tmp %>% dplyr::select(all_of(exposure_prs))))
    tsls.con = glm(as.formula(paste0('', outcome, '~ predict.con')),
                   data = tmp, family='binomial') #Fit a logistic regression model on all the participants
    f = summary(lm)$f[1]
    res <- summary(tsls.con)
    out[,1] <- outcome
    out[,2] <- exposure
    out[,3] <- c('univariate')
    out[,c(4:7)] <- res$coef[2,]
    out[,8] <- f
    names(out) <- c('outcome', 'exposure', 'model', 'b', 'se', 'zval', 'pval', 'F')
    l[[i]] <- out
    detach(tmp)
  }
  l_df <- do.call(rbind, l)
  return(l_df)
}
mr_using_prs_binary_cov <- function(exposure, exposure_prs, outcome_list, covar_list, data) {
  l <- list()
  for (i in 1:length(outcome_list)) {
    out <- as.data.frame(matrix(data = NA, nrow = 1))
    outcome <- outcome_list[i]
    print(outcome)
    tmp <- data %>% dplyr::select(all_of(c(exposure, exposure_prs, outcome_list, covar_list)))
    tmp <- na.omit(tmp)
    attach(tmp)
    con = tmp %>% dplyr::filter((!!as.name(outcome)) == 0) %>% dplyr::select(all_of(exposure_prs)) %>% unlist #values for prs in the controls only
    lm <- lm(as.formula(paste0('', exposure, ' ~ ', exposure_prs, ' + ', paste(covar_list, collapse = '+'))),
             data = tmp %>% dplyr::filter((!!as.name(outcome)) == 0))
    predict.con = predict(lm, newdata=list(con=tmp %>% dplyr::select(all_of(exposure_prs)))) #Generate predicted values for all participants based on the linear regression in the controls only.
    tsls.con = glm(as.formula(paste0('', outcome, '~ predict.con + ', paste(covar_list, collapse = '+'))),
                   data = tmp, family='binomial') #Fit a logistic regression model on all the participants
    f = summary(lm)$f[1]
    res <- summary(tsls.con)
    out[,1] <- outcome
    out[,2] <- exposure
    out[,3] <- c('univariate')
    out[,c(4:7)] <- res$coef[2,]
    out[,8] <- f
    names(out) <- c('outcome', 'exposure', 'model', 'b', 'se', 'zval', 'pval', 'F')
    l[[i]] <- out
    detach(tmp)
  }
  l_df <- do.call(rbind, l)
  return(l_df)
}
```

plot and nlmr function:

```{r}


run_nlmr_input_specific <- function(genotype,exposure,outcome, covars, family, q, logodd = F, p = 1) {
  
  tmp <- tibble(
    g = genotype,
    e = exposure,
    o = outcome
  ) %>%
    drop_na()
  
sum_data  <-create_nlmr_summary(
      y = tmp$o,
      x = tmp$e,
      g = tmp$g,
      covar = covars,
      q=q,
      family = family,
      strata_method = "residual"
    ) 


model <- with(sum_data$summary, frac_poly_summ_mr(bx=bx,
                  by=by, 
                  bxse=bxse, 
                  byse=byse, 
                  xmean=xmean,
                  family=family,
                  fig=TRUE)
)


ranked_bootstrap <- function(z) {
  seed = z + 50
sum_data2  <-create_nlmr_summary(
      y = tmp$o,
      x = tmp$e,
      g = tmp$g,
      covar = covars,
      q=q,
      seed = seed,
      family = family,
      strata_method = "ranked"
    ) 


model2 <- with(sum_data2$summary, frac_poly_summ_mr(bx=bx,
                  by=by, 
                  bxse=bxse, 
                  byse=byse, 
                  xmean=xmean,
                  family=family,
                  fig=F))


model2$lace %>%
  as_tibble() %>%
  mutate(strata = row_number()) %>%
  mutate(replicate = z) %>%
  mutate(method = "Ranked") %>%
  bind_cols(sum_data2$summary)
}

results <- map_dfr(1:p, ranked_bootstrap)
tmp2 <-model$lace %>% as_tibble() %>% mutate(method = "Residual") %>%
  mutate(strata = row_number()) %>%
  bind_cols(sum_data$summary)

tmp3 <- results %>% group_by(strata, method) %>%
  summarise(se = sqrt(sum(se^2)/n()),
            beta = mean(beta),
            bx = mean(bx),
            by = mean(by),
            bxse = sqrt(sum(bxse^2)/n()),
            byse = sqrt(sum(byse^2)/n()),
            xmean = mean(xmean)
            )%>%
  ungroup()
tmp2 %>%
  bind_rows(tmp3) %>%
         group_by(method) %>%
    ungroup() %>%
    arrange(-strata) %>%
      forestplot(name = strata,
                 estimate = beta, 
                 se = se,
                 logodds = logodd,
                 colour = method) +
      coord_flip() 
      
}


run_nlmr_w_bootstrap <- function(input,dat, covars, family, q) {
sum_data  <-create_nlmr_summary(
      y = input,
      x = dat$exposure,
      g = dat$exposure_prs,
      covar = covars,
      q=q,
      family = family,
      strata_method = "residual"
    ) 

model <- with(sum_data$summary, frac_poly_summ_mr(bx=bx,
                  by=by, 
                  bxse=bxse, 
                  byse=byse, 
                  xmean=xmean,
                  family=family,
                  fig=TRUE)
)

residual <- model$lace %>%
  as_tibble() %>%
    mutate(strata = row_number()) %>%
  mutate(method = "Residual")

ranked_bootstrap <- function(z) {
sum_data2  <-create_nlmr_summary(
      y = input,
      x = dat$exposure,
      g = dat$exposure_prs,
      seed = z,
      covar = covars,
      q=q,
      family = family,
      strata_method = "ranked"
    ) 


model2 <- with(sum_data2$summary, frac_poly_summ_mr(bx=bx,
                  by=by, 
                  bxse=bxse, 
                  byse=byse, 
                  xmean=xmean,
                  family=family,
                  fig=F))


model2$lace %>%
  as_tibble() %>%
  mutate(strata = row_number()) %>%
  mutate(replicate = z) %>%
  mutate(method = "Ranked")
}

ranked <- map_dfr(1:2, ranked_bootstrap)
ranked %>%
  group_by(strata, method) %>%
  summarise(se = sqrt(sum(se^2)/n()),
            beta = mean(beta)) %>%
  ungroup() %>%
  bind_rows(residual) %>%
  arrange(-strata)
  

}


```

Bring in data.

This has been obscured for data privacy reasons. You can use your own UK Biobank application and genetic analyses to generate this.

```{r}


dat <- vroom("~/data/non-linear/pheno/analysis_data_R1.tsv.gz")

#ensure participants have measured BMI and BMI prs

exclusions <-vroom("~/data/non-linear/geno/exclusions/data.minimal_relateds.plink.txt.gz", col_names = c("ieu", "IID"))
exclusions_highly <-vroom("~/data/non-linear/geno/exclusions/data.highly_relateds.plink.txt.gz", col_names = c("ieu", "IID"))

dat <- dat %>% 
  anti_join(exclusions) %>%
  anti_join(exclusions_highly)

```

Sub table 1:

```{r}


dat %>%
  drop_na(bmi_prs, age_at_entry) %>%

  select(sex, age_at_entry, is_dead, body_mass_index_bmi, vitd,ldl_direct, censor_time) %>%
  gtsummary::tbl_summary()
```

First, bring in Vitamin D and BMI and show they affect levels

```{r}

dat_vitd <- dat %>%
  drop_na(prs_vitd, vitd, PC1, sex, uk_biobank_assessment_centre, year_of_birth) %>%
  mutate(exposure = vitd, exposure_prs = prs_vitd) %>%
  mutate(sex = if_else(sex == "Male",1,0)) %>%
  mutate(prs_vitd = scale(prs_vitd))

dat_bmi <- dat %>%
  drop_na(body_mass_index_bmi, bmi_prs, PC1, sex, uk_biobank_assessment_centre, year_of_birth) %>%
  mutate(exposure = body_mass_index_bmi, exposure_prs = bmi_prs)%>%
  mutate(sex = if_else(sex == "Male",1,0))%>%
  mutate(bmi_prs = scale(bmi_prs))
```

Linear estimates from logistic regresion and OSMR.

```{r}

m <- lm(age_at_entry ~ bmi_prs, data = dat_bmi)
tidy(m, conf.int = T) 
m <- glm(sex ~ bmi_prs, data = dat_bmi, family = "binomial")
tidy(m, conf.int = T, exponentiate = T) %>%
  mutate(estimate = paste0(signif(estimate, 2)," (", signif(conf.low,2), "-", signif(conf.high, 2), ")"))
m <- glm(sex ~ prs_vitd, data = dat_vitd, family = "binomial")
tidy(m, conf.int = T, exponentiate = T) %>%
  mutate(estimate = paste0(signif(estimate, 3)," (", signif(conf.low,3), "-", signif(conf.high, 3), ")"))
m <- lm(age_at_entry~ prs_vitd, data = dat_vitd)
tidy(m, conf.int = T) 

m <- lm(body_mass_index_bmi ~ bmi_prs, data = dat_bmi)
glance(m)

mr_using_prs_binary(exposure = "body_mass_index_bmi", exposure_prs =  "bmi_prs", outcome_list = "sex", data =  dat_bmi) 

mr_using_prs_binary(exposure = "vitd", exposure_prs =  "vitd_prs", outcome_list = "sex", data =  dat_bmi) 


OneSampleMR::tsri(
  age_at_entry ~ body_mass_index_bmi | bmi_prs, 
  data = sleep_dat,
  link = "identity"
)

OneSampleMR::tsri(
  age_at_entry ~ vitd | prs_vitd, 
  data = sleep_dat,
  link = "identity"
)

```

Now for NCO

Run sex with covariates for Vitamin D. First, run with covars:

#VITD SEX

```{r}
dummies_no_sex <- model.matrix(~ age_at_entry +  uk_biobank_assessment_centre + PC1 + PC2 + PC3 + PC3 + PC5, data = dat_vitd)[,-1]

vitd_sex_covars <- run_nlmr_w_bootstrap(dat_vitd$sex, dat_vitd, dummies_no_sex, "binomial",10)
vitd_sex_covars_fig <- vitd_sex_covars%>%
  filter(method == "Residual") %>%
      forestplot(name = strata,
                 estimate = beta, 
                 se = se,
                 logodds = T
                 ) +
      coord_flip() +
      # geom_vline(aes(xintercept =1), col = "red", lty = "dashed") +
      ylab("Strata of Vitamin D") +
  xlab("Odds of being male")

vitd_sex_covars_fig

ggsave("vitd_on_sex.tiff", height = 6, width = 6, compression = "lzw", bg = "white")

write_rds(vitd_sex_covars_fig, "vitd_sex.rds", compress = "gz")

vitd_sex_no_covars <- run_nlmr_w_bootstrap(dat_vitd$sex, dat_vitd, NULL, "binomial",10)
vitd_sex_no_covars_fig <- vitd_sex_no_covars %>%
      forestplot(name = strata,
                 estimate = beta, 
                 se = se,
                 logodds = T,
                 colour = method) +
      coord_flip() +
      geom_vline(aes(xintercept =1), col = "red", lty = "dashed") +
      ylab("Strata of Vitamin D") +
  xlab("Odds of being male")

vitd_sex_no_covars_fig


```

#VITD AGE

```{r}
dummies_no_age <- model.matrix(~ sex +  uk_biobank_assessment_centre + PC1 + PC2 + PC3 + PC3 + PC5, data = dat_vitd)[,-1]


vitd_age_covars <- run_nlmr_w_bootstrap(dat_vitd$age_at_entry, dat_vitd, dummies_no_age, "gaussian",10)
vitd_age_no_covars <- run_nlmr_w_bootstrap(dat_vitd$age_at_entry, dat_vitd, NULL, "gaussian",10)


vitd_age_covars_fig <- vitd_age_covars %>%
      forestplot(name = strata,
                 estimate = beta, 
                 se = se,
                 logodds = F,
                 colour = method) +
      coord_flip() +
      geom_vline(aes(xintercept =0), col = "red", lty = "dashed") +
      ylab("Strata of Vitamin D") +
  xlab("Coefficent on age (years)")


vitd_age_covars_fig <- vitd_age_covars %>%
  filter(method == "Residual") %>%
      forestplot(name = strata,
                 estimate = beta, 
                 se = se,
                 logodds = F) +
      coord_flip() +
      # geom_vline(aes(xintercept =0), col = "red", lty = "dashed") +
      ylab("Strata of Vitamin D") +
  xlab("Coefficent on age (years)")

ggsave("vitd_on_age.tiff", height = 6, width = 6, compression = "lzw", bg = "white")

write_rds(vitd_age_covars_fig, "vitd_age.rds", compress = "gz")

vitd_age_no_covars_fig <- vitd_age_no_covars %>%
      forestplot(name = strata,
                 estimate = beta, 
                 se = se,
                 logodds = F,
                 colour = method) +
      coord_flip() +
      geom_vline(aes(xintercept =0), col = "red", lty = "dashed") +
      ylab("Strata of Vitamin D") +
  xlab("Coefficent on age (years)")

quantile(dat_bmi$body_mass_index_bmi, 0.99)


```

#BMI SEX

```{r}
dummies_no_sex <- model.matrix(~ age_at_entry +  uk_biobank_assessment_centre + PC1 + PC2 + PC3 + PC3 + PC5, data = dat_bmi)[,-1]

bmi_sex_covars <- run_nlmr_w_bootstrap(dat_bmi$sex, dat_bmi, dummies_no_sex, "binomial",10)

bmi_sex_covars_fig <- bmi_sex_covars %>%
      forestplot(name = strata,
                 estimate = beta, 
                 se = se,
                 logodds = T,
                 colour = method) +
      coord_flip() +
      geom_vline(aes(xintercept =1), col = "red", lty = "dashed") +
      ylab("Strata of BMI") +
  xlab("Odds ratio for male sex")


bmi_sex_no_covars <- run_nlmr_w_bootstrap(dat_bmi$sex, dat_bmi, NULL, "binomial",10)
bmi_sex_no_covars_fig <- bmi_sex_no_covars %>%
      forestplot(name = strata,
                 estimate = beta, 
                 se = se,
                 logodds = T,
                 colour = method) +
      coord_flip() +
      geom_vline(aes(xintercept =1), col = "red", lty = "dashed") +
      ylab("Strata of BMI") +
  xlab("Odds ratio for male sex")


bmi_sex_covars_fig


```

#BMI AGE

```{r}
dummies_no_age <- model.matrix(~ sex +  uk_biobank_assessment_centre + PC1 + PC2 + PC3 + PC3 + PC5, data = dat_bmi)[,-1]


bmi_age_covars <- run_nlmr_w_bootstrap(dat_bmi$age_at_entry, dat_bmi, dummies_no_age, "gaussian",10)
bmi_age_no_covars <- run_nlmr_w_bootstrap(dat_bmi$age_at_entry, dat_bmi, NULL, "gaussian",10)
bmi_age_covars_fig <- bmi_age_covars %>%
      forestplot(name = strata,
                 estimate = beta, 
                 se = se,
                 logodds = F,
                 colour = method) +
      coord_flip() +
      geom_vline(aes(xintercept =0), col = "red", lty = "dashed") +
      ylab("Strata of BMI") +
  xlab("Coefficent on age (years)")


bmi_age_no_covars_fig <- bmi_age_no_covars %>%
      forestplot(name = strata,
                 estimate = beta, 
                 se = se,
                 logodds = F,
                 colour = method) +
      coord_flip() +
      geom_vline(aes(xintercept =0), col = "red", lty = "dashed") +
      ylab("Strata of BMI") +
  xlab("Coefficent on age (years)")


bmi_age_covars_fig

```

#PLOTS

```{r}

Fig1 <- (vitd_age_covars_fig + vitd_sex_covars_fig) /
(bmi_age_covars_fig + bmi_sex_covars_fig)  + plot_annotation(tag_levels = "A")+ plot_layout(guides = "collect") & theme(legend.position = 'bottom')

Fig1
ggsave("Fig1.tiff", device = "tiff", compression = "lzw", plot = Fig1, bg= "white")

FigS1 <- (vitd_age_no_covars_fig + vitd_sex_no_covars_fig) /
(bmi_age_no_covars_fig + bmi_sex_no_covars_fig)  + plot_annotation(tag_levels = "A")+ plot_layout(guides = "collect") & theme(legend.position = 'bottom')

FigS1

```

#VITD AGE + Sex - logged

```{r}

dat_vitd_log <- dat_vitd %>%
  mutate(exposure = log(vitd))

vitd_log_age_no_covars<- run_nlmr_w_bootstrap(dat_vitd_log$age_at_entry, dat_vitd_log, NULL, "gaussian",10)
vitd_log_age_no_covars_fig <- vitd_log_age_no_covars %>%
      forestplot(name = strata,
                 estimate = beta, 
                 se = se,
                 logodds = F,
                 colour = method) +
      coord_flip() +
      geom_vline(aes(xintercept =0), col = "red", lty = "dashed") +
      ylab("Strata of Vitamin D") +
  xlab("Coefficent on age (years)")


vitd_log_sex_no_covars<- run_nlmr_w_bootstrap(dat_vitd_log$sex, dat_vitd_log, NULL, "binomial",10)
vitd_log_sex_no_covars_fig <- vitd_log_sex_no_covars %>%
      forestplot(name = strata,
                 estimate = beta, 
                 se = se,
                 logodds = T,
                 colour = method) +
      coord_flip() +
      geom_vline(aes(xintercept =1), col = "red", lty = "dashed") +
      ylab("Strata of Vitamin D") +
  xlab("Odds of being male")

vitd_log_sex_no_covars_fig / vitd_log_age_no_covars_fig + plot_annotation(tag_levels = "A") + plot_layout(guides = "collect") & theme(legend.position = 'bottom')
  
  
```

#smoking BMI

```{r}
dat_bmi_smoke <- dat_bmi %>%
  filter(ever_smoked == "Yes")
dat_bmi_no_smoke <- dat_bmi %>%
  filter(ever_smoked == "No")

bmi_age_smoke <- run_nlmr_w_bootstrap(dat_bmi_smoke$age_at_entry, dat_bmi_smoke, NULL, "gaussian",10)
bmi_age_no_smoke <- run_nlmr_w_bootstrap(dat_bmi_no_smoke$age_at_entry, dat_bmi_no_smoke, NULL, "gaussian",10)
bmi_sex_smoke <- run_nlmr_w_bootstrap(dat_bmi_smoke$sex, dat_bmi_smoke, NULL, "binomial",10)
bmi_sex_no_smoke <- run_nlmr_w_bootstrap(dat_bmi_no_smoke$sex, dat_bmi_no_smoke, NULL, "binomial",10)


bmi_age_smoke_fig <- bmi_age_smoke %>%
      forestplot(name = strata,
                 estimate = beta, 
                 se = se,
                 logodds = F,
                 colour = method) +
      coord_flip() +
      geom_vline(aes(xintercept =0), col = "red", lty = "dashed") +
      ylab("Strata of BMI (smokers)") +
  xlab("Coefficent on age (years)")


bmi_age_no_smoke_fig <- bmi_age_no_smoke %>%
      forestplot(name = strata,
                 estimate = beta, 
                 se = se,
                 logodds = F,
                 colour = method) +
      coord_flip() +
      geom_vline(aes(xintercept =0), col = "red", lty = "dashed") +
      ylab("Strata of BMI (non-smokers)") +
  xlab("Coefficent on age (years)")

bmi_sex_no_smoke_fig <- bmi_sex_no_smoke %>%
      forestplot(name = strata,
                 estimate = beta, 
                 se = se,
                 logodds = T,
                 colour = method) +
      coord_flip() +
      geom_vline(aes(xintercept =1), col = "red", lty = "dashed") +
      ylab("Strata of BMI (non-smokers)") +
  xlab("Odds of being male")

bmi_sex_smoke_fig <- bmi_sex_smoke %>%
      forestplot(name = strata,
                 estimate = beta, 
                 se = se,
                 logodds = T,
                 colour = method) +
      coord_flip() +
      geom_vline(aes(xintercept =1), col = "red", lty = "dashed") +
      ylab("Strata of BMI (smokers)") +
  xlab("Odds of being male")

(bmi_sex_no_smoke_fig + bmi_sex_smoke_fig) / (bmi_age_no_smoke_fig + bmi_age_smoke_fig) + plot_annotation(tag_levels = "A") + plot_layout(guides = "collect") & theme(legend.position = 'bottom')

```

#smoking Vit D

```{r}
dat_vitd_smoke <- dat_vitd %>%
  filter(ever_smoked == "Yes")
dat_vitd_no_smoke <- dat_vitd %>%
  filter(ever_smoked == "No")

vitd_age_smoke <- run_nlmr_w_bootstrap(dat_vitd_smoke$age_at_entry, dat_vitd_smoke, NULL, "gaussian",10)
vitd_age_no_smoke <- run_nlmr_w_bootstrap(dat_vitd_no_smoke$age_at_entry, dat_vitd_no_smoke, NULL, "gaussian",10)
vitd_sex_smoke <- run_nlmr_w_bootstrap(dat_vitd_smoke$sex, dat_vitd_smoke, NULL, "binomial",10)
vitd_sex_no_smoke <- run_nlmr_w_bootstrap(dat_vitd_no_smoke$sex, dat_vitd_no_smoke, NULL, "binomial",10)


vitd_age_smoke_fig <- vitd_age_smoke %>%
      forestplot(name = strata,
                 estimate = beta, 
                 se = se,
                 logodds = F,
                 colour = method) +
      coord_flip() +
      geom_vline(aes(xintercept =0), col = "red", lty = "dashed") +
      ylab("Strata of Vitamin D (smokers)") +
  xlab("Coefficent on age (years)")


vitd_age_no_smoke_fig <- vitd_age_no_smoke %>%
      forestplot(name = strata,
                 estimate = beta, 
                 se = se,
                 logodds = F,
                 colour = method) +
      coord_flip() +
      geom_vline(aes(xintercept =0), col = "red", lty = "dashed") +
      ylab("Strata of Vitamin D (non-smokers)") +
  xlab("Coefficent on age (years)")

vitd_sex_no_smoke_fig <- vitd_sex_no_smoke %>%
      forestplot(name = strata,
                 estimate = beta, 
                 se = se,
                 logodds = T,
                 colour = method) +
      coord_flip() +
      geom_vline(aes(xintercept =1), col = "red", lty = "dashed") +
      ylab("Strata of Vitamin D (non-smokers)") +
  xlab("Odds of being male")

vitd_sex_smoke_fig <- vitd_sex_smoke %>%
      forestplot(name = strata,
                 estimate = beta, 
                 se = se,
                 logodds = T,
                 colour = method) +
      coord_flip() +
      geom_vline(aes(xintercept =1), col = "red", lty = "dashed") +
      ylab("Strata of Vitamin D (smokers)") +
  xlab("Odds of being male")

(vitd_sex_no_smoke_fig + vitd_sex_smoke_fig) / (vitd_age_no_smoke_fig + vitd_age_smoke_fig) + plot_annotation(tag_levels = "A") + plot_layout(guides = "collect") & theme(legend.position = 'bottom')

```

#Other non-lin outcomes

```{r}
other_outcomes <- vroom("~/data/non-linear/breast_feeding.tsv.gz")

other <- dat %>%
  left_join(other_outcomes) %>%
  rename(mat_smok = p1787_i0, north = p129_i0, breastfed = p1677_i0, birthweight = p20022_i0)

other %>%
  count(breastfed)
other <- other %>%

  mutate(mat_smok = if_else(str_detect(mat_smok, "Yes|No"),  mat_smok, NA_character_)) %>%
  mutate(mat_smok = mat_smok=="Yes") %>%
   mutate(breastfed = if_else(str_detect(breastfed, "Yes|No"),  breastfed, NA_character_)) %>%
  mutate(breastfed = breastfed=="Yes") %>%
  mutate(north = as.numeric(north)/1000) %>%
  mutate(prs_vitd = scale(prs_vitd), bmi_prs = scale(bmi_prs))

tbl <- bind_rows(
tidy(glm(mat_smok ~bmi_prs, data = other, family = "binomial")),
tidy(glm(breastfed ~bmi_prs, data = other, family = "binomial")),
tidy(lm(birthweight ~bmi_prs, data = other)),
tidy(lm(north ~bmi_prs, data = other)),
)%>%
  filter(term == "bmi_prs") %>%
  mutate(outcome = c("Maternal smoking", "Breastfeeding", "Birthweight (Kg)", "Latitude (Km)")) %>%
  mutate(IV = "BMI")


p.breastfed <- run_nlmr_input_specific(other$bmi_prs,
                        other$body_mass_index_bmi,
                        other$breastfed, 
                        covars = NULL,
                        family = "binomial",
                        logodd = T,
                        q = 10) +
  geom_vline(aes(xintercept =1), col = "red", lty = "dashed")  +
  xlab("Odds Ratio (breastfeeding)") +
  ylab("Strata of BMI")


p.maternal_smoke <- run_nlmr_input_specific(other$bmi_prs,
                        other$body_mass_index_bmi,
                        other$breastfed, 
                        covars = NULL,
                        family = "binomial",
                        logodd = T,
                        q = 10) +
  geom_vline(aes(xintercept =1), col = "red", lty = "dashed")  +
  xlab("Odds ratio (maternal smoking)") +
  ylab("Strata of BMI")

p.birthweight <- run_nlmr_input_specific(other$bmi_prs,
                        other$body_mass_index_bmi,
                        other$birthweight, 
                        covars = NULL,
                        family = "gaussian",
                        logodd = F,
                        q = 10) +
  geom_vline(aes(xintercept =0), col = "red", lty = "dashed")  +
  xlab("beta on birthweight (kg)") +
  ylab("Strata of BMI")


p.north <- run_nlmr_input_specific(other$bmi_prs,
                        other$body_mass_index_bmi,
                        other$north/1000, 
                        covars = NULL,
                        family = "gaussian",
                        logodd = F,
                        q = 10) +
  geom_vline(aes(xintercept =0), col = "red", lty = "dashed")  +
  xlab("beta on latitude (km)") +
  ylab("Strata of BMI")
p_bmi <- (p.birthweight + p.north) / (p.maternal_smoke + p.breastfed) +
  plot_annotation(tag_levels = "A") +
  plot_layout(guides = "collect") & theme(legend.position = "bottom") 

p_bmi
```

other non lin Now for vit D

```{r}
tbl2 <- bind_rows(
tidy(glm(mat_smok ~prs_vitd, data = other, family = "binomial")),
tidy(glm(breastfed ~prs_vitd, data = other, family = "binomial")),
tidy(lm(birthweight ~prs_vitd, data = other)),
tidy(lm(north/1000 ~prs_vitd, data = other)),
) %>%
    filter(term == "prs_vitd") %>%
  mutate(outcome = c("Maternal smoking", "Breastfeeding", "Birthweight (Kg)", "Latitude (Km)")) %>%
  mutate(IV = "Vitamin D")


tbl %>%
  bind_rows(tbl2) %>%
  select(-statistic) %>%
  pivot_wider(id_cols = c(outcome), names_from = term, values_from = c(estimate, p.value, std.error)) %>%
  relocate(outcome, estimate_bmi_prs, std.error_bmi_prs, p.value_bmi_prs, estimate_prs_vitd, std.error_prs_vitd) %>%
  mutate(across(is.numeric, ~ signif(.x,3))) %>%
  gt::gt()
p.breastfed <- run_nlmr_input_specific(other$prs_vitd,
                        other$vitd,
                        other$breastfed, 
                        covars = NULL,
                        family = "binomial",
                        logodd = T,
                        q = 10) +
  geom_vline(aes(xintercept =1), col = "red", lty = "dashed")  +
  xlab("Odds Ratio (breastfeeding)") +
  ylab("Strata of Vitamin D")
p.maternal_smoke <- run_nlmr_input_specific(other$prs_vitd,
                        other$vitd,
                        other$breastfed, 
                        covars = NULL,
                        family = "binomial",
                        logodd = T,
                        q = 10) +
  geom_vline(aes(xintercept =1), col = "red", lty = "dashed")  +
  xlab("Odds ratio (maternal smoking)") +
  ylab("Strata of Vitamin D")
p.birthweight <- run_nlmr_input_specific(other$prs_vitd,
                        other$vitd,
                        other$birthweight, 
                        covars = NULL,
                        family = "gaussian",
                        logodd = F,
                        q = 10) +
  geom_vline(aes(xintercept =0), col = "red", lty = "dashed")  +
  xlab("beta on birthweight (kg)") +
  ylab("Strata of Vitamin D")


p.north <- run_nlmr_input_specific(other$prs_vitd,
                        other$vitd,
                        other$north/1000, 
                        covars = NULL,
                        family = "gaussian",
                        logodd = F,
                        q = 10) +
  geom_vline(aes(xintercept =0), col = "red", lty = "dashed")  +
  xlab("beta on latitude (km)") +
  ylab("Strata of Vitamin D")
p_vitd <- (p.birthweight + p.north) / (p.maternal_smoke + p.breastfed) +
  plot_annotation(tag_levels = "A") +
  plot_layout(guides = "collect") & theme(legend.position = "bottom") 

p_vitd 
```

DL-

```{r}

LDL_dat <- dat %>%
  drop_na(ldl_direct, sex, age_at_entry, prs_LDL) %>%
  mutate(sex = sex=="Male") %>%
  mutate(prs_LDL = scale(prs_LDL))


LDL_age_osmr <- OneSampleMR::tsri(
  age_at_entry ~ ldl_direct | prs_LDL, 
  data = LDL_dat,
  link = "identity"
)

LDL_age_osmr
LDL_sex_osmr <- OneSampleMR::tsri(
  sex ~ ldl_direct | prs_LDL, 
  data = LDL_dat,
  link = "logit"
)

LDL_sex_osmr

```

Figure 3

```{r}

LDL_on_sex <- run_nlmr_input_specific(LDL_dat$prs_LDL, LDL_dat$ldl_direct, LDL_dat$sex, covars = NULL, family = "binomial", q = 10, logodd = T, p = 2)

LDL_on_sex <- LDL_on_sex +
  xlab("OR of being male") +
  ylab("Strata of LDL")


LDL_on_age <- run_nlmr_input_specific(LDL_dat$prs_LDL, LDL_dat$ldl_direct, LDL_dat$age_at_entry, covars = NULL, family = "gaussian", q = 10, logodd = F, p = 2)

LDL_on_age <- LDL_on_age +
  xlab("beta on age") +
  ylab("Strata of LDL")


LDL_on_age + LDL_on_sex + plot_layout(guides = "collect") + plot_annotation(tag_levels = "A")& theme(legend.position = 'bottom')
```

ON MI

```{r}
LDL_future_mi_osmr <- OneSampleMR::tsri(
  future_MI~ ldl_direct | prs_LDL, 
  data = LDL_dat,
  link = "logit"
)

LDL_ever_mi_osmr <- OneSampleMR::tsri(
  ever_MI~ ldl_direct | prs_LDL, 
  data = LDL_dat,
  link = "logit"
)

LDL_dead_osmr <- OneSampleMR::tsri(
  is_dead~ ldl_direct | prs_LDL, 
  data = LDL_dat,
  link = "logit"
)

LDL_dead_osmr
```

Figure 4

```{r}


LDL_on_history_MI <- run_nlmr_input_specific(LDL_dat$prs_LDL, LDL_dat$ldl_direct, LDL_dat$history_MI, covars = NULL, family = "binomial", q = 10, logodd = T)

LDL_on_history_MI


LDL_on_future_MI <- run_nlmr_input_specific(LDL_dat$prs_LDL, LDL_dat$ldl_direct, LDL_dat$future_MI, covars = NULL, family = "binomial", q = 10, logodd = T)

LDL_on_future_MI


LDL_on_ever_MI <- run_nlmr_input_specific(LDL_dat$prs_LDL, LDL_dat$ldl_direct, LDL_dat$ever_MI, covars = NULL, family = "binomial", q = 10, logodd = T)

LDL_on_ever_MI <- LDL_on_ever_MI +
  xlab("OR for myocardial infarction") +
  ylab("Strata of LDL")


LDL_on_death <- run_nlmr_input_specific(LDL_dat$prs_LDL, LDL_dat$ldl_direct, LDL_dat$is_dead, covars = NULL, family = "binomial", q = 10, logodd = T)

LDL_on_death <- LDL_on_death +
  xlab("OR for death") +
  ylab("Strata of LDL")


LDL_on_ever_MI + LDL_on_death + plot_layout(guides = "collect") + plot_annotation(tag_levels = "A") & theme(legend.position = 'bottom')


```

Supplementary Figures 4 and 5

```{r}
LDL_on_history_MI <- LDL_on_history_MI +
  xlab("OR for prevalent myocardial infarction") +
  ylab("Strata of LDL")


LDL_on_future_MI <- LDL_on_future_MI +
  xlab("OR for incident myocardial infarction") +
  ylab("Strata of LDL")

LDL_on_history_MI + LDL_on_future_MI + plot_layout(guides = "collect") + plot_annotation(tag_levels = "A") & theme(legend.position = 'bottom')
```

```{r}

LDL_on_death <- run_nlmr_input_specific(LDL_dat$prs_LDL, LDL_dat$ldl_direct, LDL_dat$is_dead, covars = NULL, family = "binomial", q = 100, logodd = T)


```

With covars (supplementary)

```{r}

dummies_LDL <- model.matrix(~ age_at_entry + sex + uk_biobank_assessment_centre + PC1 + PC2 + PC3 + PC3 + PC5,  data = LDL_dat)[,-1]


LDL_on_history_MI_covar <- run_nlmr_input_specific(LDL_dat$prs_LDL, LDL_dat$ldl_direct, LDL_dat$history_MI, covars = dummies_LDL, family = "binomial", q = 10, logodd = T)

LDL_on_history_MI


LDL_on_future_MI_covar <- run_nlmr_input_specific(LDL_dat$prs_LDL, LDL_dat$ldl_direct, LDL_dat$future_MI, covars = dummies_LDL, family = "binomial", q = 10, logodd = T)

LDL_on_future_MI_covar$data


LDL_on_ever_MI_covar <- run_nlmr_input_specific(LDL_dat$prs_LDL, LDL_dat$ldl_direct, LDL_dat$ever_MI, covars = dummies_LDL, family = "binomial", q = 10, logodd = T, p = 1)

LDL_on_ever_MI_covar <- LDL_on_ever_MI_covar + 
  xlab("OR for myocardial infarction") +
  ylab("Strata of LDL")

LDL_on_dead_covar <- run_nlmr_input_specific(LDL_dat$prs_LDL, LDL_dat$ldl_direct, LDL_dat$is_dead, covars = dummies_LDL, family = "binomial", q = 10, logodd = T)

LDL_on_dead_covar <- LDL_on_dead_covar + 
  xlab("OR for death") +
  ylab("Strata of LDL")
LDL_on_ever_MI_covar + LDL_on_dead_covar + plot_layout(guides = "collect") + plot_annotation(tag_levels = "A") & theme(legend.position = 'bottom')

```

Figure 5

```{r}

sum_data <- SUMnlmr::create_nlmr_summary(g = LDL_dat$prs_LDL,
                             x = LDL_dat$ldl_direct,
                             y = LDL_dat$is_dead,
                             covar = NULL,
                             family = "binomial",q = 100,
                             strata_method = "ranked"
                            
                            )


model<- with(sum_data$summary, frac_poly_summ_mr(bx=bx,
                  by=by, 
                  bxse=bxse, 
                  byse=byse, 
                  xmean=xmean,
                  family="binomial",
                  fig=TRUE, )
)
model$figure +
  xlab("LDL-C (mmol/L)") +
  ylab("OR of death with increasing LDL-C")

model$lace
```

Add in statins

```{r}
statin <- fread("STATIN_FILE") 

LDL_dat <- LDL_dat %>%
  mutate(statin = if_else(eid %in% statin$V2, 1,0)) 


dummies_LDL_statin <- model.matrix(~ age_at_entry + sex + statin + uk_biobank_assessment_centre + PC1 + PC2 + PC3 + PC3 + PC5, data = LDL_dat)[,-1]


LDL_on_ever_MI_covar_statin <- run_nlmr_input_specific(LDL_dat$prs_LDL, LDL_dat$ldl_direct, LDL_dat$ever_MI, covars = dummies_LDL_statin, family = "binomial", q = 10, logodd = T)

p_statin_adj <- LDL_on_ever_MI_covar_statin +
  xlab("OR of ever MI") +
  ylab("Strata of LDL (adjusted for statins)")

LDL_dat %>%
  mutate(age_c = ntile(age_at_entry,10)) %>%
  group_by(age_c) %>%
  summarise(stat = mean(statin), age = mean(age_at_entry))

LDL_dat %>%
  filter(age_at_entry < 50) %>%
  summarise(stat = mean(statin), age = mean(age_at_entry))

LDL_young <- LDL_dat %>%
  filter(age_at_entry <50) 

LDL_statin <- LDL_dat %>%
  filter(statin  == 1)

LDL_nostatin <- LDL_dat %>%
  filter(statin  == 0)


```

```{r}

LDL_on_ever_MI_young <- run_nlmr_input_specific(LDL_young$prs_LDL, LDL_young$ldl_direct, LDL_young$ever_MI, covars = NULL, family = "binomial", q = 10, logodd = T)

p_young <- LDL_on_ever_MI_young +
    xlab("OR of ever MI") +
  ylab("Strata of LDL (< 50 only)")

LDL_on_ever_MI_nostatin <- run_nlmr_input_specific(LDL_nostatin$prs_LDL, LDL_nostatin$ldl_direct, LDL_nostatin$ever_MI, covars = NULL, family = "binomial", q = 10, logodd = T)

p_statin_non_user <- LDL_on_ever_MI_nostatin +
    xlab("OR of ever MI") +
  ylab("Strata of LDL (non-statin users only)")

LDL_on_ever_MI_statin <- run_nlmr_input_specific(LDL_statin$prs_LDL, LDL_statin$ldl_direct, LDL_statin$ever_MI, covars = NULL, family = "binomial", q = 10, logodd = T)

p_statin_user <- LDL_on_ever_MI_statin +
    xlab("OR of ever MI") +
  ylab("Strata of LDL (statin users only)")


(p_statin_adj + p_young) / (p_statin_user + p_statin_non_user) + plot_layout(guides = "collect") + plot_annotation(tag_levels = "A") & theme(legend.position = 'bottom')
```

```{r}
LDL_on_dead_statin <- run_nlmr_input_specific(LDL_statin$prs_LDL, LDL_statin$ldl_direct, LDL_statin$is_dead, covars = NULL, family = "binomial", q = 10, logodd = T)

LDL_on_dead_nostatin <- run_nlmr_input_specific(LDL_nostatin$prs_LDL, LDL_nostatin$ldl_direct, LDL_nostatin$is_dead, covars = NULL, family = "binomial", q = 10, logodd = T)

LDL_on_dead_young <- run_nlmr_input_specific(LDL_young$prs_LDL, LDL_young$ldl_direct, LDL_young$is_dead, covars = NULL, family = "binomial", q = 10, logodd = T)


LDL_on_statin  <- run_nlmr_input_specific(LDL_dat$prs_LDL, LDL_dat$ldl_direct, LDL_dat$statin, covars = NULL, family = "binomial", q = 10, logodd = T)

LDL_on_statin


```

Other bits of code.

```{r}
tg_dat <- vroom("LDL/ldl_tg.csv")
df <- tg_dat %>%
mutate(OR_CI = `Odds ratio/Hazard ratio`) %>%
  mutate(
    numeric_values = gsub("[^0-9. ]", "", OR_CI),
    values = strsplit(numeric_values, "\\s+"),
    OR = map_dbl(values, function(x) as.numeric(x[1])),
    CI_lower = map_dbl(values, function(x) as.numeric(x[2])),
    CI_upper = map_dbl(values, function(x) as.numeric(x[3]))
  ) %>%
  select(-numeric_values, -values)


# Calculate beta and SE
df <- df %>%
  mutate(
    beta = log(OR),
    SE = (log(CI_upper) - log(CI_lower)) / 3.96)
  )
  
  
df %>%
  filter(Outcome == "Cancer mortality") %>%
  arrange(-Stratum) %>%
  forestplot(name = Stratum,
             estimate = beta,
             se = SE) +
  coord_flip() +
  xlab("Odds of cancer mortality") +
  ylab("Strata of TG")
```

```{r}

tg_dat <- vroom("LDL/ldl_cad.csv")
df <- tg_dat %>%
mutate(OR_CI = `Odds ratio/Hazard ratio`) %>%
  mutate(
    numeric_values = gsub("[^0-9. ]", "", OR_CI),
    values = strsplit(numeric_values, "\\s+"),
    OR = map_dbl(values, function(x) as.numeric(x[1])),
    CI_lower = map_dbl(values, function(x) as.numeric(x[2])),
    CI_upper = map_dbl(values, function(x) as.numeric(x[3]))
  ) %>%
  select(-numeric_values, -values)


# Calculate beta and SE
df <- df %>%
  mutate(
    beta = log(OR),
    se = (log(CI_upper) - log(CI_lower)) / 3.96) %>%
  mutate(method = "Yang et al") %>%
  mutate(strata = factor(Stratum, ordered = T))
  )
  
df2 <- LDL_on_ever_MI_covar$data %>%
  filter(method == "Ranked") %>%
  mutate(Stratum = as.double(strata))

tidy(lm(beta ~ Stratum, weights = se, data = df2), conf.int = T)

exp()
df %>%
  select(strata, Stratum)
LDL_on_ever_MI_covar$data %>%
  filter(method == "Ranked") %>%
  mutate(method = "This paper") %>%
  mutate(beta = log(beta)) %>%
  bind_rows(df) %>%
  forestplot(estimate = beta, se = se, colour = method, name = strata, logodds = T) +
  coord_flip() +
  theme(legend.position = "bottom") +
  xlab("Odds ratio of MI (ours) /CAD (Yang et al)") +
  ylab("Strata")

LDL_on_ever_MI_covar$data %>%
  filter(method == "Ranked") %>%
  mutate(beta = log(beta)) %>% 

LDL_on_ever_MI_covar$data %>%
  filter(method == "Ranked") %>%
  select(xmean, beta, se)

LDL_on_ever_MI_covar

LDL_on_ever_MI_covar <- run_nlmr_input_specific(LDL_dat$prs_LDL, LDL_dat$ldl_direct, LDL_dat$ever_MI, covars = dummies_LDL, family = "binomial", q = 10, logodd = T, p = 1)


df2 %>%
  bind_rows(df) %>%
  mutate(xmean = if_else(is.na(xmean), `mean-ldl`, xmean)) %>%
  select(xmean, method, strata) %>%
  pivot_wider(values_from = xmean, names_from = method) %>%
  ggplot(aes(x = Ranked, y = Yours)) +
  geom_point() +
  geom_smooth(method = "lm") +
  xlab("Our strata mean LDL") +
  ylab("Your strata mean LDL")

dat$bmi_prs
```

Further plots in discussion including TG and alluvial plots

```{r}
{r}

dat_tg <- dat %>%
  drop_na(prs_TG, tg1, cancer_death)


dummies <- model.matrix(~ age_at_entry + age_at_entry^2 + sex + age_at_entry* sex + age_at_entry^2 * sex + uk_biobank_assessment_centre + PC1 + PC2 + PC3 + PC3 + PC5 + PC6 + PC7 + PC8 +PC9 + PC10, data = dat_tg)[,-1]

TG_cancer_osmr <- OneSampleMR::tsri(
  cancer_death ~ tg2 | prs_TG, 
  data = dat_tg,
  link = "logit"
)

TG_cancer_osmr
tg_cancer_tsmr <- run_nlmr_input_specific(dat$prs_TG, dat$tg1, dat$cancer_death, covars = dummies, family = "binomial", q = 10, logodd = T)

p1 <- tg_cancer_tsmr +
  xlab("OR for cancer mortality per mmol/L") + 
  ylab("Strata") + scale_color_manual(values=c("#F8766D", "#00BFC4")) +
  theme(legend.position = "bottom")

p1
ggsave("tg_cancer.svg", bg = "white", height = 6, width = 8)
tg_cancer_tsmr$data %>%
  filter(method == "Ranked") %>%
  select(Stratum = strata, xmean) %>%
  mutate(Stratum = as.numeric(Stratum)) %>%
  left_join(tg_dat) %>%
  summarise(cor = cor(TG, xmean, use = "complete.obs"))

dat_tg %>%
  drop_na(tg1, tg2) %>%
  summarise(cor = cor(tg1, tg2))

p2 <- dat_tg %>%
  drop_na(tg2) %>%
  mutate(tri1 = generate_ranked_strata(prs_TG, tg1, 10),
         tri2 = generate_ranked_strata(prs_TG, tg2, 10)) %>%
  count(tri1,tri2) %>%
  filter(tri1 == 1 | tri1 == 2) %>%
  as_data_frame() %>%
  drop_na() %>%
  ggplot(aes(y = n, axis1 = tri1, axis2 = tri2, fill = tri1)) +
  geom_alluvium( width = 1/12) +
  geom_stratum(width = 1/12, fill = "black", color = "grey") +
  geom_label(stat = "stratum", aes(label = after_stat(stratum))) +
  scale_x_discrete(limits = c("UKB study entry", "Repeat Sample"), expand = c(.05, .05)) +
  ylab("Number of participants") +
  xlab("Triglyceride measurements over two time points") +

  theme_bw() +
  theme(legend.position = "none") 

p3 <- dat_tg %>%
  drop_na(tg2) %>%
  mutate(tri1 = generate_ranked_strata(prs_TG, tg1, 10),
         tri2 = generate_ranked_strata(prs_TG, tg2, 10)) %>%
  count(tri1,tri2) %>%
  # filter(tri1 == 1 | tri1 == 2) %>%
  as_data_frame() %>%
  drop_na() %>%
  ggplot(aes(y = n, axis1 = tri1, axis2 = tri2, fill = tri1)) +
  geom_alluvium( width = 1/12) +
  geom_stratum(width = 1/12, fill = "black", color = "grey") +
  geom_label(stat = "stratum", aes(label = after_stat(stratum))) +
  scale_x_discrete(limits = c("UKB study entry", "Repeat Sample"), expand = c(.05, .05)) +
  ylab("Number of participants") +
  xlab("Triglyceride measurements over two time points") +

  theme_bw() +
  theme(legend.position = "none") 

p2
p3
ggsave("tg_variable_all.svg", bg = "white", height = 12, width = 8)
dat %>%
  count(cancer_death)

dat_tg %>%
  drop_na(tg2) %>%
  mutate(tri1 = generate_ranked_strata(prs_TG, tg1, 10),
         tri2 = generate_ranked_strata(prs_TG, tg2, 10)) %>%
  count(tri1,tri2) %>%
  filter(tri1 == 1 | tri1 == 2) %>%
  as_data_frame() %>%
  drop_na()  %>%
  group_by(tri1) %>%
  mutate(perc = n/sum(n))
```
````
